# Dense Longitudinal Multi-Omics Reveals Microbial–Metabolic–Immune Succession During Immune Checkpoint Therapy

**DOI:** 10.64898/2026.10.24.26366392

**Authors:** Ebrahim Zandi, Kwat Medetgul-Ernar, Heinz-Joseph Lenz, Farshad Alishahi

## Abstract

Immune checkpoint inhibitors (ICIs) can induce durable clinical responses, yet relationships among the gut microbiome, microbial function, intestinal metabolism, and host immunity remain incompletely resolved. We performed dense longitudinal multi-omics profiling of a single patient with colorectal cancer metastatic to the liver during ICI therapy and sustained tumor regression. Eleven serial stool samples spanning approximately 7.3 months were analyzed by 16S rRNA sequencing, microbial metatranscriptomics, metabolomics, lipidomics, and stool host-RNA immune profiling and aligned with serial clinical imaging. A major intestinal ecosystem transition accompanied the emergence of clinical response. Microbial abundance and transcriptional activity were frequently discordant, demonstrating that compositional profiling alone incompletely captured microbial dynamics. Taxon-resolved functional analysis revealed sequential microbial states rather than a single responder-associated configuration. Loss of early Bacteroides/Fusobacterium-associated functions accompanied purine/sphingolipid remodeling and changes in myeloid-associated immune signatures. At T5, a transient Bifidobacterium carbohydrate/fermentation program coincided with lactate/indole-3-lactate remodeling and broad adaptive T-cell, memory/regulatory, mucosal, and B-cell immune changes. At T6, marked Clostridium transcriptional emergence and lipid/coenzyme-A pathway activation coincided with fatty-acid/lysophospholipid remodeling and a distinct immune configuration. Historical HUMAnN reconstruction using original UniRef90 and MetaCyc resources confirmed taxon-specific, reaction-associated gene-family support for prioritized functional programs. These findings define a temporally ordered microbial–metabolic–immune succession during ICI response. Although this N-of-1 design cannot establish population generalizability, dense within-person multi-omics resolves transient functional states obscured by cross-sectional analyses and generates testable hypotheses for larger prospective cohorts and personalized studies of ICI response.

## Introduction

Immune checkpoint blockade has shown substantial antitumor efficacy activity in microsatellite instability-high (MSI-H), mismatch repair-deficient (dMMR) metastatic colorectal cancer, including durable responses to PD-1 inhibition alone or in combination with CTLA-4 blockade [1, 2]. Nevertheless, treatment responses remain heterogeneous, highlighting the importance of understanding additional host and environmental factors that may influence therapeutic response. Among these, the intestinal microbiome has emerged as a potentially modifiable contributor to antitumor immunity and immune checkpoint inhibitor (ICI) efficacy [3–6]. Early clinical studies identified differences in pretreatment gut microbial communities between ICI responders and nonresponders [3–5], while interventional studies demonstrated that fecal microbiota transplantation (FMT) from ICI-responsive donors can restore sensitivity to anti-PD-1 therapy in a subset of refractory patients, accompanied by immune remodeling [7, 8]. Higher dietary fiber intake has also been associated with improved progression-free survival during ICI therapy [9].

The mechanisms connecting intestinal microorganisms to antitumor immunity extend beyond bacterial abundance. Microorganisms generate or modify metabolites capable of influencing immune cell differentiation and function. For example, microbiome-derived inosine can enhance checkpoint blockade under appropriate experimental conditions [10], whereas *Fusobacterium nucleatum*-derived succinate has been implicated in impaired antitumor T-cell trafficking and resistance to immunotherapy in colorectal cancers [11]. Nevertheless, individual microbial taxa associated with ICI response have shown limited reproducibility across cohorts [12]. This heterogeneity may reflect differences in patient populations, treatment regimens, environmental exposures, and microbial functional redundancy. Different organisms can perform overlapping metabolic functions, while individual taxa may occupy distinct transcriptional states depending on ecological and host context. Consequently, taxonomic abundance alone may incompletely represent the biologically active microbial ecosystem. Characterizing microbial functional activity and its associated chemical environment may therefore provide additional insight into microbiome-ICI interactions.

Most human microbiome-ICI studies have emphasized pretreatment sampling, providing limited information about how intestinal microbial communities evolve during treatment. Recent longitudinal studies in melanoma and primary liver cancer demonstrate that microbial taxa and functional pathways can undergo treatment-associated changes that are not captured by baseline measurements alone [13–16]. Although large cohorts are essential for identifying reproducible associations and establishing population generalizability, dense within-individual multi-omics integration addresses a complementary question: how biological changes unfold over time within a single patient. By using each individual as a longitudinal reference, repeated sampling can reduce the influence of between-person heterogeneity, resolve transient or sequential molecular states, and identify temporal relationships among independently measured biological compartments [15, 17–19]. Such analyses can generate candidate mechanisms for subsequent testing in larger cohorts.

In the present study, we applied dense longitudinal multi-omics integration to 11 serial stool specimens collected over approximately 7.3 months from a patient with MSI-H/dMMR colorectal cancer metastatic to the liver who experienced sustained tumor regression during nivolumab plus ipilimumab therapy. We integrated 16S rRNA sequencing, microbial metatranscriptomics, stool metabolomics, lipidomics, and host-derived immune transcriptional signatures to characterize longitudinal changes in the intestinal ecosystem. By distinguishing microbial abundance from transcriptional activity and reconstructing historical HUMAnN pathway-to-reaction-to-UniRef90 relationships, we sought to identify molecularly supported microbial functional programs and their temporal associations with measured chemicals and host immune states. Our objective was to determine whether dense longitudinal integration could resolve distinct stages of intestinal ecosystem remodeling and generate testable microbial-metabolic-immune hypotheses for validation in larger prospective ICI cohorts.

## Methods

### Study design, clinical course, and longitudinal sampling

Eleven serial stool specimens (T0-T10) were collected over approximately 7.3 months at the USC Norris Comprehensive Cancer Center from a patient with microsatellite instability-high (MSI-H), mismatch repair-deficient (dMMR) colorectal cancer metastatic to the liver. The patient received combination immune checkpoint blockade with nivolumab (anti-PD-1) and ipilimumab (anti-CTLA-4) for treatment of the liver metastasis. The first stool specimen (T0) was collected 17 days before treatment initiation. Nivolumab was administered intravenously at approximately 3 mg/kg every two weeks (16 administrations), with ipilimumab administered concurrently at approximately 1 mg/kg every six weeks (six administrations). Individual doses were adjusted according to body weight. The complete de-identified treatment schedule is provided in Supplementary_Treatment_Table. The complete 16S and microbial taxon-activity matrices are provided in Supplementary Tables S2 and S3.

Serial computed tomography (CT) imaging were used to assess changes in the dominant hepatic lesion, with tumor burden estimated from comparable coronal cross-sectional areas normalized to baseline (100%). To preserve patient confidentiality, calendar dates were replaced with elapsed months relative to the first stool collection (T0 = month 0). For descriptive analyses, the molecular series was divided into early/pre-transition (T0-T4), transition (T5-T6), and later/post-transition (T7-T10) periods, with T5 and T6 examined separately in temporal-mechanistic analyses. Microbial, metabolomic, and lipidomic measurements extended through T10, whereas continuous stool host-RNA immune measurements were available through T9.

### Ethics approval and institutional protocol

This study was conducted under the institutional research protocol entitled *“The Role of Microbiome in Cancer Therapy”* (Protocol No. OS-16-9), with oversight by the University of Southern California Institutional Review Board (USC IRB No. HS-CG-13-0003). Longitudinal clinical and multi-omics data were analyzed using de-identified study identifiers, with calendar dates replaced by relative time from the first stool collection to protect patient confidentiality. Written informed consent was obtained from the participant in accordance with the approved institutional protocol.

### 16S rRNA and metatranscriptomic profiling

Bacterial community structure was characterized via 16S rRNA gene sequencing. DNA extraction was performed by bead beating using Qiagen Powersoil kits.Amplification of the V4 hypervariable region of the 16S rRNA gene were performed according to the published protocol [20, 21]. The library underwent 2×250 sequencing on an Illumina HiSeq 2500 to a mean depth of 107,433 merged sequences per sample. QIIME 1.9.1 was used to perform quality filtering, merge paired end reads, and cluster sequences into 97% operational taxonomic units [22]. Taxonomy was assigned using the Greengenes May 2013 database [23].

Genus-level 16S entries were aggregated after taxonomic suffix harmonization, and unresolved assignments were excluded. The 15 most variable genera were selected by longitudinal standard deviation independently of clinical, chemical, or immune data and standardized within genus (z-scores) for visualization.

### Stool metabolomic and lipidomic profiling

Longitudinal stool metabolomic and lipidomic analyses were performed at the West Coast Metabolomics Center, University of California, Davis (Fiehn Laboratory). The facility’s metabolomics workflows included gas chromatography-time-of-flight mass spectrometry (GC-TOF MS) for primary metabolites and hydrophilic interaction liquid chromatography coupled to electrospray quadrupole time-of-flight tandem mass spectrometry (HILIC-ESI-QTOF MS/MS) for biogenic amines and other polar metabolites. Metabolite identification involved chromatographic retention characteristics, mass-spectral library matching, and quality-control procedures, with abundances reported as relative semiquantitative measurements.

Untargeted lipidomic profiling was performed using reverse-phase liquid chromatography coupled to high-resolution mass spectrometry (LC-HRMS). Approximately 4mg of each stool specimen was extracted using the Matyash method [24] with methyl tert-butyl ether, methanol, and water. The organic phase was collected, dried, and reconstituted in methanol/toluene (9:1, v/v) containing 50 ng/mL CUDA. Lipid extracts were separated using an Agilent 1290 Infinity LC system equipped with a Waters ACQUITY UPLC CSH C18 column and analyzed in positive-and negative-electrospray ionization modes using Agilent 6550 and 6530 quadrupole time-of-flight mass spectrometers, respectively.

The longitudinal metabolomics dataset comprised 246 annotated metabolites across T0-T10, together with individual lipid species representing multiple lipid classes, including ceramides, sphingomyelins (SM), lysophosphatidylcholines (LPC), and phosphatidylethanolamines (PE). The lipidomics data were provided by the university of California Davis core as internal-standard-normalized relative intensities. The 40 most longitudinally variable metabolites were selected using a scale-aware variability measure and standardized within feature for heatmap visualization; individual metabolite trajectories retained the core-provided quantitative measurements. Lipid-class trajectories were calculated as means of individually standardized constituent species, whereas ceramide/SM and LPC/PE ratios were calculated from summed internal-standard-normalized intensities before longitudinal standardization. Chemical features were analyzed independently of clinical tumor response before integration with prioritized microbial functional pathways. Additional analytical parameters and computational procedures are provided in Supplementary Methods.

### Stool RNA extraction, library preparation, and sequencing

Stool samples were stored at −80°C until processing. Total RNA was extracted using the Norgen Stool Total RNA Purification Kit, with DNase treatment performed both on-column and after extraction. RNA concentration, purity, and integrity were assessed using Qubit RNA High Sensitivity, NanoDrop, and TapeStation, respectively. Host and bacterial ribosomal RNA was depleted using QIAseq FastSelect rRNA/globin and FastSelect-5S/16S/23S reagents. Strand-specific sequencing libraries were prepared using the low-RNA-integrity-number (low-RIN) input protocol of the NEBNext Ultra II Directional RNA Library Prep Kit with NEB index adapters. Libraries were pooled at equimolar concentrations and sequenced on an Illumina NovaSeq X Plus platform using 2×150-bp paired-end reads, targeting approximately 50 million read pairs per sample.

### Human transcriptomic processing and immune-feature profiling

Raw RNA sequencing data were assessed using FastQC v0.12.1 [25] and preprocessed with fastp v1.0.1 [26]. Preprocessing included automatic paired-end adapter detection, poly-X/poly-G trimming, Phred quality filtering (Q20), removal of reads containing more than 40% low-quality bases, a minimum retained read length of 36 bp, and additional 3′-end trimming.

Human transcripts were quantified using kallisto v0.52.0 [27] against a combined Ensembl GRCh38 cDNA and noncoding RNA reference, indexed with a k-mer length of 21 [28]. Pseudoalignment accounted for the reverse-forward strandedness of the libraries. Transcript abundances were expressed as transcripts per million (TPM), aggregated into gene-level scores, and log-transformed. Gene-set enrichment scores were generated using Molecular Signatures Database (MSigDB) [29] and Caucell Inc. gene sets (Caucell Inc., Stanford, CA, USA) and immune-component scores were calculated using the Caucell Inc. immune models.

The core facility evaluated associations between gene-, gene-set-, and immune-component scores and the clinical response conditions using information correlation-based analyses [30, 31]. Features with an absolute correlation coefficient ≥0.8 or adjusted P-value ≤0.05 were included in the core’s exploratory association tables. Immune profiles were visualized using longitudinal plots, heatmaps, bubble plots, Immune GPS, and Immunograms.

For the present longitudinal analyses, 12 RNA-derived immune population/signature scores were evaluated across T0-T9 (Fig. 4). Higher-resolution analyses used 65 continuous immune features spanning eight immune models: overall immune, granulocyte, monocyte, natural-killer, T-invariant, CD8 T-cell, CD4 T-cell, and B-cell systems (Figs. 4 and 5; Supplementary Figs. S7 and S8). Continuous immune scores were used for quantitative longitudinal and multi-omic integration, whereas percentile/rank-transformed Bubble and Immunogram representations were retained for visualization and quality control. These measurements were interpreted as immune-associated transcriptional signatures rather than direct immune-cell counts.

### Microbial metatranscriptomic processing and functional profiling

Microbial transcriptional activity was analyzed separately from the human transcriptomic data using the sequencing core’s microbial bioinformatics workflow. Read preprocessing and host-sequence depletion were performed using KneadData v0.7.8.1 with Trimmomatic v0.33 [32] and the human hg39 T2T Bowtie2 reference [32]. Microbial taxonomic profiling used MetaPhlAn with the mpa_vJan25_CHOCOPhlAnSGB_202503 database [33]. Gene-family and pathway profiling were performed using HUMAnN v3.9 with ChocoPhlAn v201901_v31, UniRef90 2019_01, and MetaCyc v24 [34, 35].

Taxon-stratified UniRef90 gene-family counts per million (CPM) were used to estimate microbial transcriptional activity, while taxon-stratified MetaCyc profiles characterized expressed microbial functional programs. Genus-level taxonomic assignments were harmonized with the 16S dataset to compare microbial abundance and transcriptional activity. The original taxon-stratified gene-family measurements were retained for subsequent molecular verification.

### Integrated microbial functional, chemical, and immune analysis

Multi-omic integration followed a sequential workflow designed to minimize circular feature selection. Microbial taxa were first prioritized using genus-harmonized 16S relative abundance and metatranscriptomic activity, independently of chemical, immune, and clinical-response measurements. Taxon-stratified MetaCyc [35] pathways were then evaluated to identify emerging and declining functional programs.

Biologically supported pathways were integrated with measured stool metabolites and lipids using longitudinal Pearson and Spearman correlations, first-difference correlations, and directional concordance across T4→T5, T5→T6, and T4→T6. Redundant pathway–chemical relationships were consolidated into five primary functional axes representing Clostridium lipid/coenzyme A (CoA) activity, Bifidobacterium fermentation, Bifidobacterium carbohydrate metabolism, Fusobacterium functional loss, and loss of the early Bacteroides/Fusobacterium functional state.

Molecular support for prioritized pathways was evaluated by reconstructing the historical MetaCyc v24 pathway-to-reaction-to-UniRef90 [36] relationships using the core’s structured pathway definitions and metacyc_reactions_level4ec_only.uniref mapping resource. Corresponding UniRef90 families were intersected with taxon-stratified transcripts detected in the patient’s original HUMAnN data [33]. Molecular support for microbial pathways was distinguished from direct biochemical attribution of measured metabolites or lipids.

The five predefined functional axes were subsequently integrated with the 65 continuous host immune features. Associations were evaluated using longitudinal and first-difference correlations and transition-specific changes. All analyses were interpreted descriptively within the single-subject longitudinal design; temporal associations were not considered evidence of causality.

### Statistical considerations & computational environment

All analyses represent serial observations from a single participant (N=1). Correlations, first-difference associations, fold changes, and z-scores describe within-person temporal structure and are not population-level inferential statistics. Analyses used Python (pandas, NumPy, SciPy, Matplotlib). Full longitudinal matrices, pathway/reaction/UniRef90 mappings, functional-axis definitions, and figure-driving audit tables should accompany the manuscript as Supplementary Data.

Large language models (including OpenAI ChatGPT and Google Gemini) assisted with code drafting, debugging, and language editing. The authors retain responsibility for independently reviewing and validating analytical procedures, results, and manuscript content. Analysis scripts and source/audit tables will be made available upon request.

## Results

### Longitudinal clinical response establishes the biological framework

Serial clinical imaging demonstrated progressive regression of the hepatic metastasis during nivolumab plus ipilimumab therapy, providing the clinical context for interpreting the longitudinal multi-omic measurements (Fig. 1A,B and Supplementary Fig. S1). The first stool specimen (T0) was collected 17 days before treatment initiation, establishing a pretreatment reference. Nivolumab was administered every two weeks for 16 documented infusions, with ipilimumab administered concurrently every six weeks for six infusions (Fig. 1A; Supplementary Table S1). The 11 longitudinal stool specimens (T0-T10) spanned the early treatment period, the emergence of radiographic response, and subsequent sustained tumor regression.

**Figure 1.**
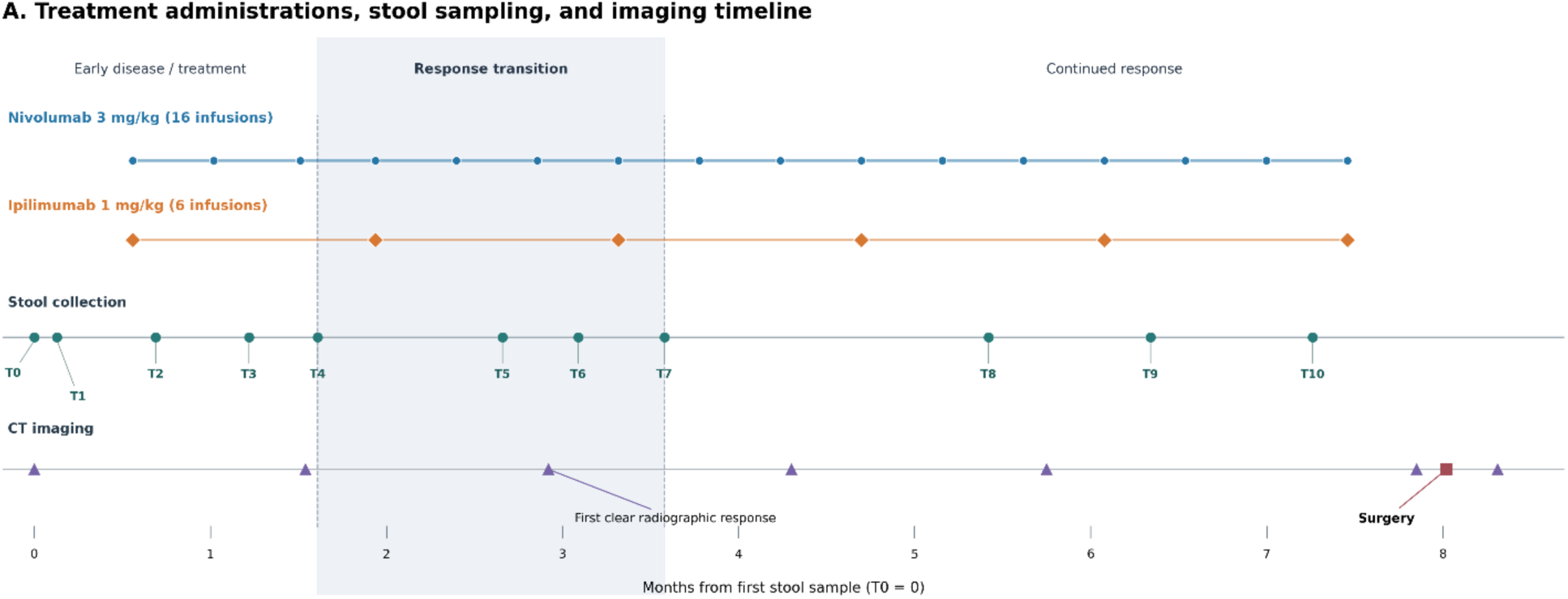
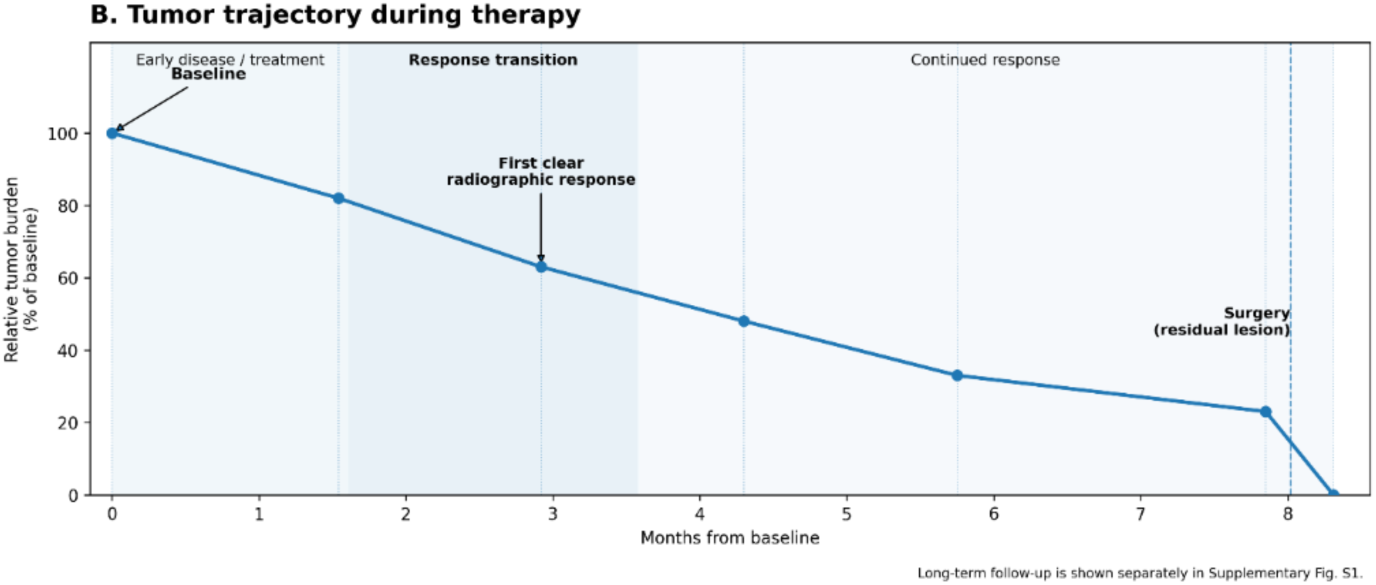
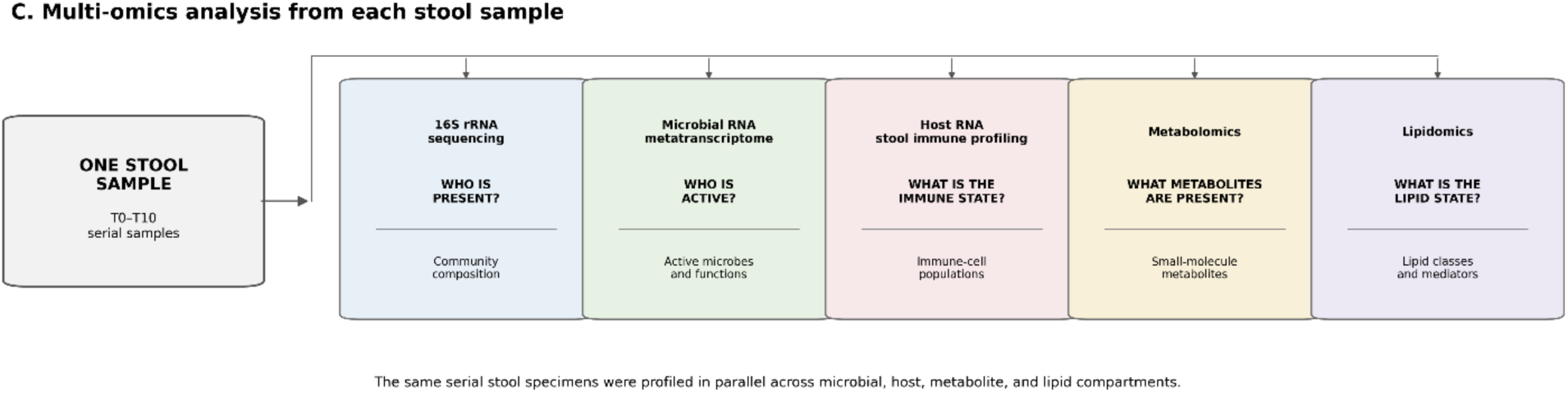
Clinical response and longitudinal multi-omics study design. (A) Treatment administration, stool sampling, and clinical imaging timeline. Eleven longitudinal stool samples (T0–T10) were collected over approximately 7.3 months during treatment of colorectal cancer metastatic to the liver with combined immune checkpoint blockade. The first stool specimen (T0) was collected 17 days before treatment initiation. Nivolumab (anti-PD-1; approximately 3 mg/kg) was administered every two weeks for a total of 16 infusions, while ipilimumab (anti-CTLA-4; approximately 1 mg/kg) was administered concurrently every six weeks for a total of six infusions. Individual administrations of nivolumab (blue circles) and ipilimumab (orange diamonds) are shown alongside stool collections (teal circles), serial computed tomography (CT) examinations (purple triangles), and surgical resection of the residual liver lesion (red square). To preserve patient confidentiality, calendar dates are omitted, and all events are expressed as months relative to the first stool collection (T0 = month 0). The clinical course is divided into early disease/treatment, response-transition, and continued-response intervals. The first clear radiographic response was observed at approximately month 2.9, during the interval encompassing the major multi-omics transition. Long-term follow-up imaging, obtained approximately 74.6 months after baseline, is presented separately in Supplementary Figure S1. (B) Longitudinal hepatic lesion response. The relative cross-sectional area of the dominant hepatic lesion was estimated from comparable serial CT images and expressed as a percentage of the baseline measurement (100%). The lesion area progressively decreased to approximately 82%, 63%, 48%, 33%, and 23% of baseline. The final value of 0% represents the post-resection state and does not indicate complete spontaneous radiographic disappearance of the lesion. The residual lesion was subsequently surgically resected; therefore, the final value of 0% denotes the post-resection state rather than an imaging-derived measurement demonstrating spontaneous disappearance of the lesion. The first clear radiographic response and subsequent period of continued tumor regression are indicated. As in panel A, time is expressed as months from baseline rather than calendar dates. (C) Longitudinal stool multi-omics design. Serial stool specimens collected from the same patient were analyzed in parallel using 16S rRNA sequencing to characterize microbial community composition; microbial RNA metatranscriptomics to identify transcriptionally active organisms and microbial functions; host RNA profiling to characterize the intestinal immune-cell state; metabolomics to measure small-molecule metabolites; and lipidomics to characterize lipid classes and mediators. This matched longitudinal design enabled microbial composition, microbial activity, host immune responses, metabolites, and lipids to be examined within the same biological specimens throughout therapy. Together, these data establish the temporal framework for subsequent analyses of coordinated microbial, functional, metabolic, lipid, and host immune changes accompanying the transition from early disease to sustained therapeutic response.

During the early treatment period, substantial tumor burden persisted, with the first follow-up CT showing a relative lesion area of approximately 82% of baseline. By approximately month 2.9, the relative area had decreased to 63%, representing the first clear radiographic evidence of a major response (Fig. 1B). Tumor regression continued thereafter, reaching 48% at month 4.3, 33% at month 5.8, and 23% at approximately month 7.9 (Fig. 1B). The residual hepatic lesion was subsequently surgically resected; therefore, the final 0% value represents the postoperative state rather than complete spontaneous radiographic disappearance. Long-term follow-up imaging obtained approximately 74.6 months after baseline is shown in Supplementary Figure S1.

Importantly, the first clear radiographic response occurred during the interval encompassing the major intestinal ecosystem transition identified in subsequent multi-omic analyses. Stool specimens were available immediately before, during, and after this period, enabling temporal comparison of microbial composition and transcriptional activity, metabolomic and lipidomic profiles, and host-derived immune signatures (Fig. 1C). For descriptive analyses, the longitudinal series was divided into early (T0-T4), transition (T5-T6), and later (T7 onward) intervals, while individual time periods were retained for temporal-mechanistic integration. These intervals represent successive observations within one patient rather than independent biological groups.

Together, the clinical timeline and matched longitudinal specimens establish the framework for examining intestinal microbial, chemical, and host immune remodeling during a sustained response to combination checkpoint blockade.

### A major intestinal microbial transition coincided with the emergence of clinical response

Longitudinal 16S rRNA sequencing and microbial metatranscriptomics revealed major restructuring of the intestinal ecosystem during the interval associated with clinical response (Fig. 2). For descriptive comparison, the series was divided into pre-transition (T0-T4; approximately months 0-1.6), transition (T5-T6; approximately months 2.7-3.1), and post-transition (T7-T10; approximately months 3.6-7.3) states.

**Figure 2.**
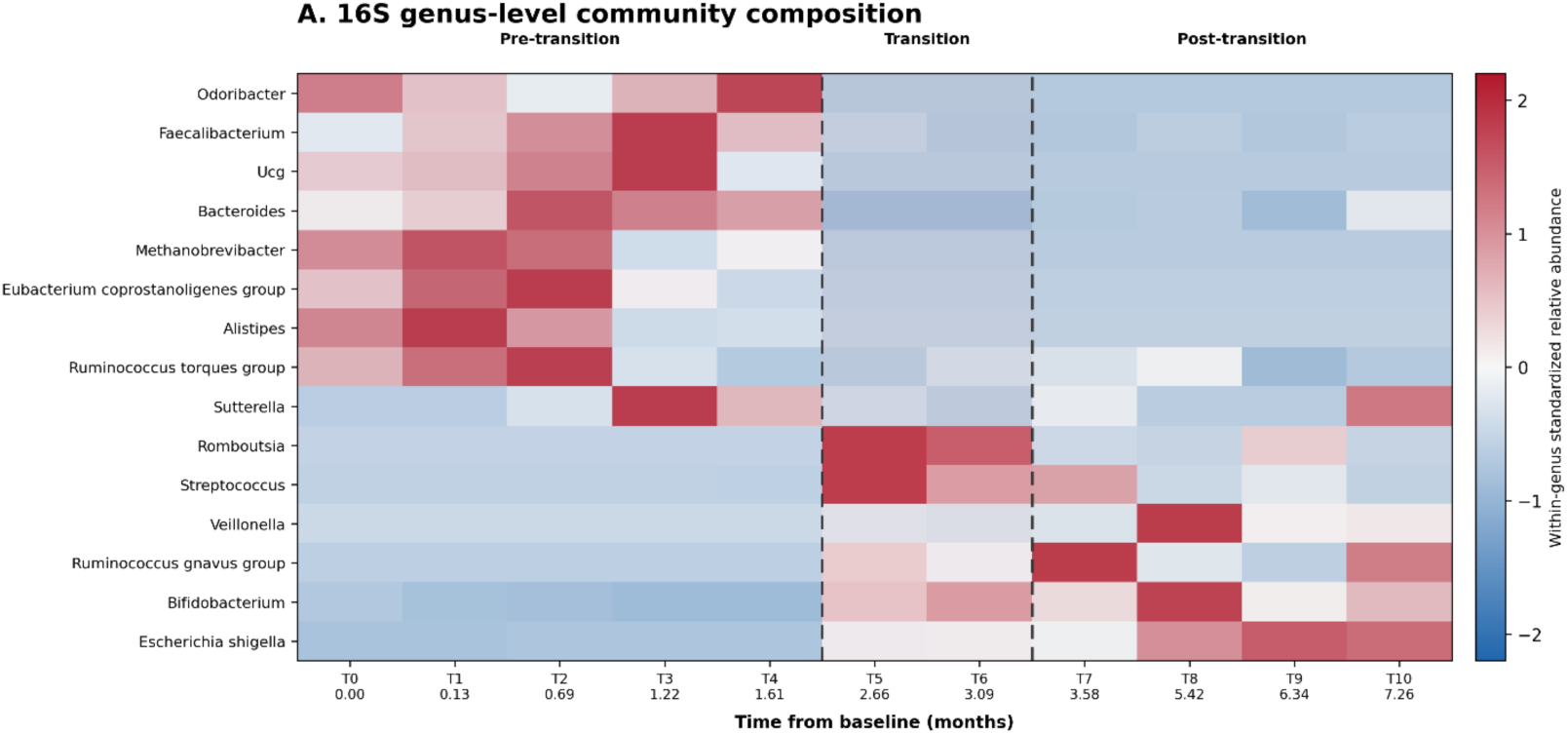
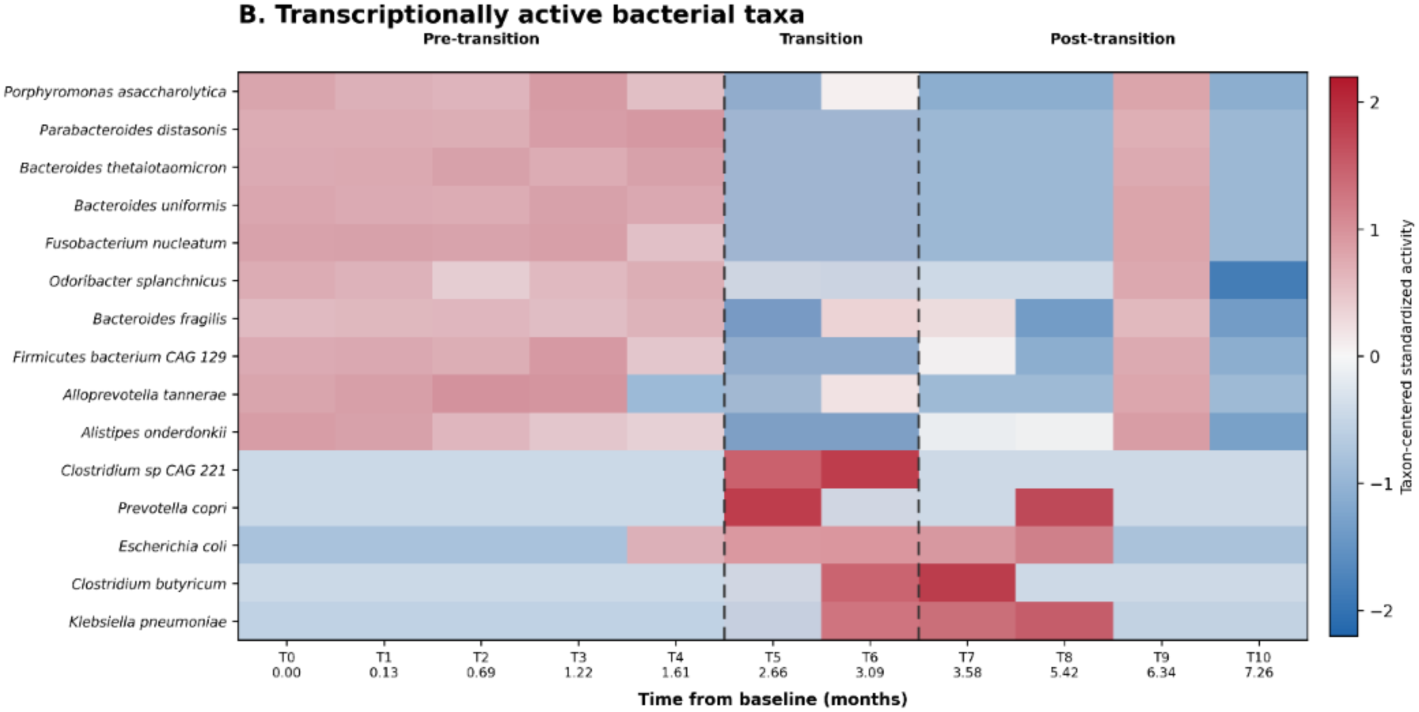
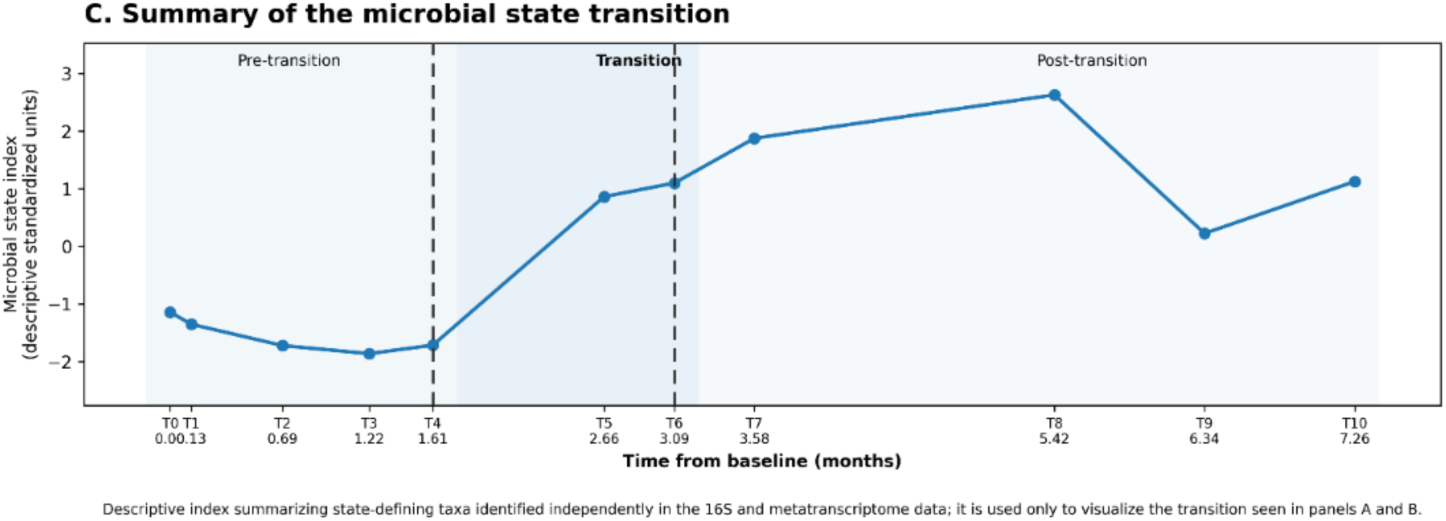
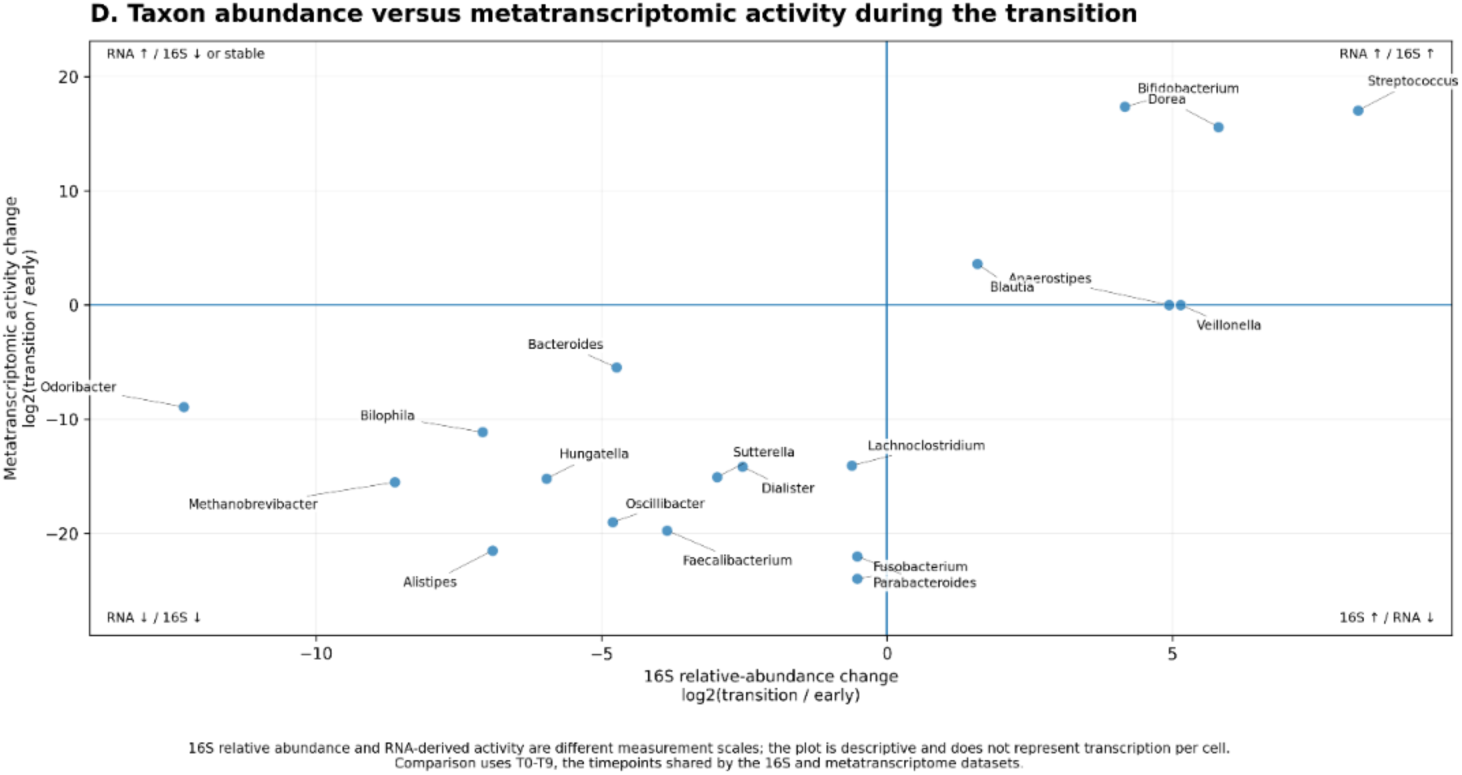
Longitudinal restructuring of intestinal microbial composition and transcriptional activity during immune checkpoint therapy (A) Longitudinal genus-level intestinal microbial community composition. The heatmap shows the 15 microbial genus-level features exhibiting the greatest longitudinal variability in 16S rRNA relative abundance across T0–T10, including the archaeal genus *Methanobrevibacter*. Taxonomic entries resolving to the same genus were aggregated before analysis. Each genus was independently standardized across the longitudinal series (z-score); therefore, colors represent relative changes within each genus rather than absolute abundance differences between genera. Samples are displayed chronologically, with elapsed months from the first stool collection shown beneath each timepoint. Dashed vertical lines delineate the early (T0–T4), transition (T5–T6), and later (T7–T10) intervals. The transition was characterized by declining early-state signals from genera including *Odoribacter*, *Alistipes*, and *Faecalibacterium*, together with increased relative abundance of selected later-state genera, including *Bifidobacterium*, *Streptococcus*, and *Veillonella*. These changes describe compositional restructuring and do not necessarily reflect corresponding changes in microbial transcriptional activity. (B) Longitudinal transcriptional activity of bacterial taxa. Heatmap shows the 15 bacterial taxa making the largest contributions to the taxon-stratified microbial transcript pool derived from metatranscriptomic sequencing. Transcriptional activity was quantified from taxon-stratified UniRef90 counts per million (CPM) and standardized independently within each taxon across the longitudinal series. Thus, colors indicate changes in taxon-attributed RNA activity over time and do not represent absolute transcript abundance between taxa. The metatranscriptomic transition is characterized by pronounced loss of activity from several early transcriptionally active taxa, including *Bacteroides*, *Fusobacterium*, *Odoribacter*, and *Alistipes*, together with emergence of a distinct later transcriptional state that includes marked activation of *Clostridium* and other transition/post-transition taxa. Samples are shown chronologically using the same temporal boundaries as in panel A. (C) Descriptive summary of the longitudinal microbial-state transition. A microbial-state index was constructed independently from the 16S and metatranscriptomic datasets by identifying features showing the strongest early-to-later state changes within each data layer, calculating standardized early-and later-state signatures, and combining the resulting layer-specific indices. Negative values therefore indicate greater similarity to the early microbial state and positive values indicate increasing similarity to the later state. The index shows a pronounced transition beginning around T5–T6 and continuing into the post-transition interval. This index is a descriptive summary of patterns independently evident in panels A and B and was not optimized against tumor burden, clinical response, metabolomic, lipidomic, or immune measurements. (D) Genus-harmonized comparison of bacterial abundance and metatranscriptomic activity during the transition. Scatter plot compares the change in genus-level 16S relative abundance (x-axis) with the corresponding change in taxon-attributed metatranscriptomic activity (y-axis), expressed as log2 transition-to-early ratios. Taxonomic assignments from the 16S and metatranscriptomic datasets were harmonized at genus level before comparison. Horizontal and vertical zero lines divide taxa according to the direction of compositional and transcriptional change. The upper-right quadrant identifies taxa showing coordinated increases in abundance and RNA activity, including *Bifidobacterium* and *Streptococcus*, whereas taxa in the lower-left quadrant undergo coordinated compositional and transcriptional loss. Importantly, taxa exhibiting disproportionately large transcriptional changes relative to their compositional changes provide evidence of abundance–activity discordance: some organisms undergo marked transcriptional activation with little corresponding increase in 16S abundance, whereas others lose transcriptional activity disproportionately to their compositional change. *Clostridium* provides a prominent example of transcriptional emergence despite limited compositional expansion, while *Fusobacterium* exhibits pronounced transcriptional deactivation relative to its change in abundance. These abundance–activity relationships provided the unbiased starting point for the taxon-resolved functional analyses in Figure 5. For all panels, sampling time is expressed as months relative to the first stool collection (T0 = month 0), and samples remain in their true chronological order. The pre-transition, transition, and post-transition designations are descriptive temporal intervals rather than independent biological groups. 16S relative abundance and metatranscriptomic RNA activity represent distinct measurement scales; RNA activity should not be interpreted as transcription per bacterial cell. Together, the four panels demonstrate that the treatment-associated intestinal microbial transition involved both community compositional restructuring and extensive redistribution of microbial transcriptional activity, with substantial abundance–activity discordance that could not be resolved from taxonomic profiling alone.

Genus-level 16S profiles showed prominent early signals from *Odoribacter, Alistipes*, and *Faecalibacterium*, followed by their decline and increases in other genera, including *Streptococcus*, *Bifidobacterium*, and *Escherichia/Shigella* at later timepoints (Fig. 2A; Supplementary Figs. S2A and S3). Community composition continued to vary after the initial transition rather than settling into a single static configuration.

Taxon-stratified metatranscriptomics revealed a parallel but nonidentical redistribution of the active microbial transcript pool (Fig. 2B). Early contributions from *Bacteroides*, *Fusobacterium*, *Odoribacter*, and related anaerobes declined, while *Prevotella*, *Escherichia*, and later *Clostridium*-associated activity increased at different timepoints (Supplementary Fig. S2B). Genus-harmonized comparisons demonstrated substantial abundance–activity discordance (Fig. 2D): Clostridium showed marked RNA emergence despite relatively limited compositional expansion, whereas Fusobacterium showed pronounced transcriptional loss relative to its 16S change. Thus, composition alone did not fully capture functional restructuring. Directional genus-harmonized activation, loss, and restructuring rankings are provided in Supplementary Table S5.

A descriptive microbial-state index summarized the shift from the early community toward the later configuration (Fig. 2C; Supplementary Table S4). The transition remained evident under alternative temporal boundaries (Supplementary Fig. S4). These findings motivated the subsequent taxon-resolved functional analysis without using tumor response to rank taxa.

### The microbial transition was accompanied by a major metabolic-state change

Stool metabolomic and lipidomic measurements showed broad chemical remodeling during the same longitudinal interval as the microbial transition (Fig. 3). Global metabolomic profiling demonstrated substantial longitudinal reorganization across the treatment course (Fig. 3A). The 40 most longitudinally variable, outcome-independent metabolites revealed distinct shifts across purine metabolism, tryptophan and indole pathways, carbohydrate-derived sugar acids, amino acid metabolism, and central carbon metabolism. This pattern reflected widespread metabolic restructuring across the complete metabolome (Supplementary Fig. S5; Supplementary Tables S6 and S8).

**Figure 3.**
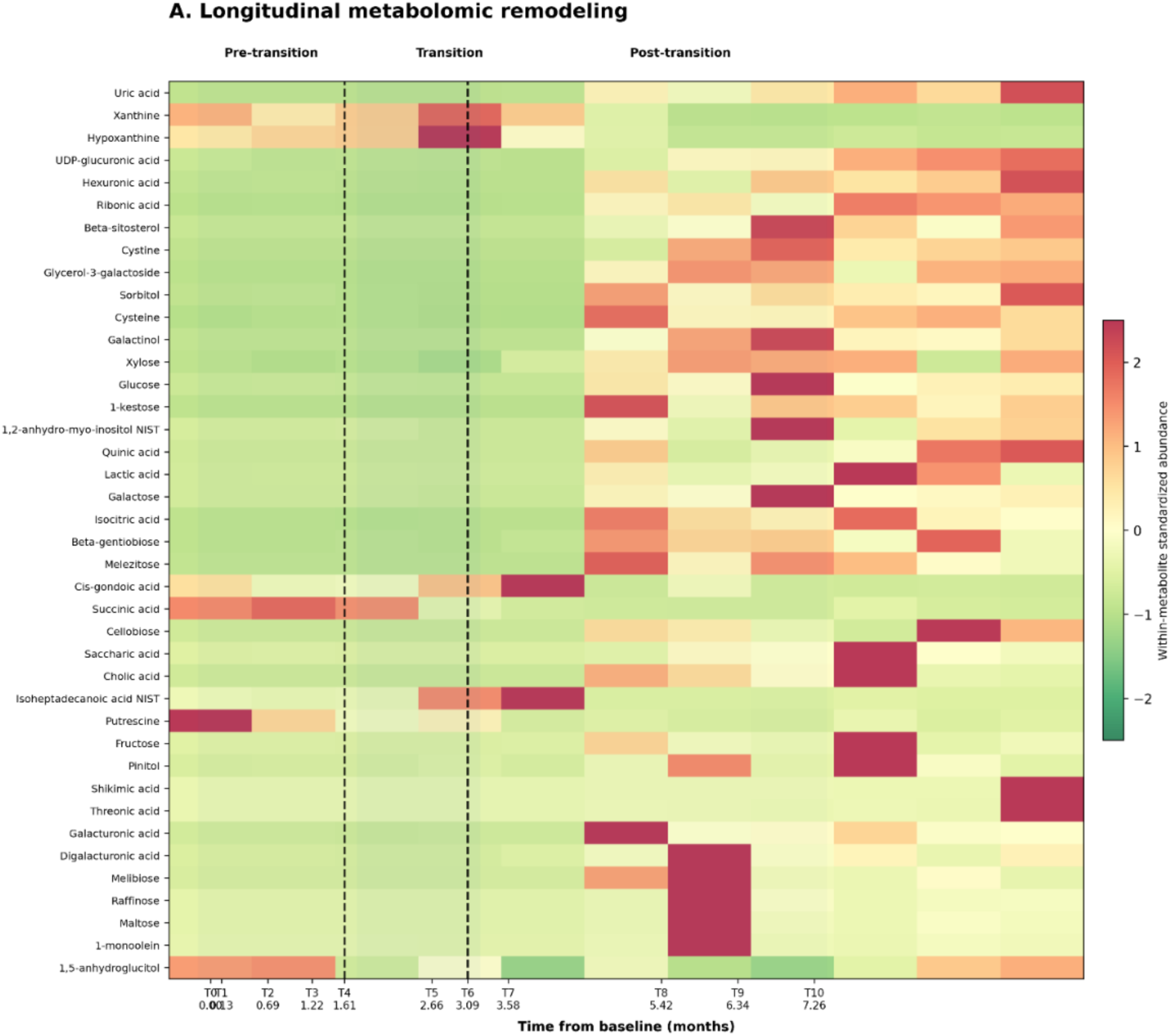
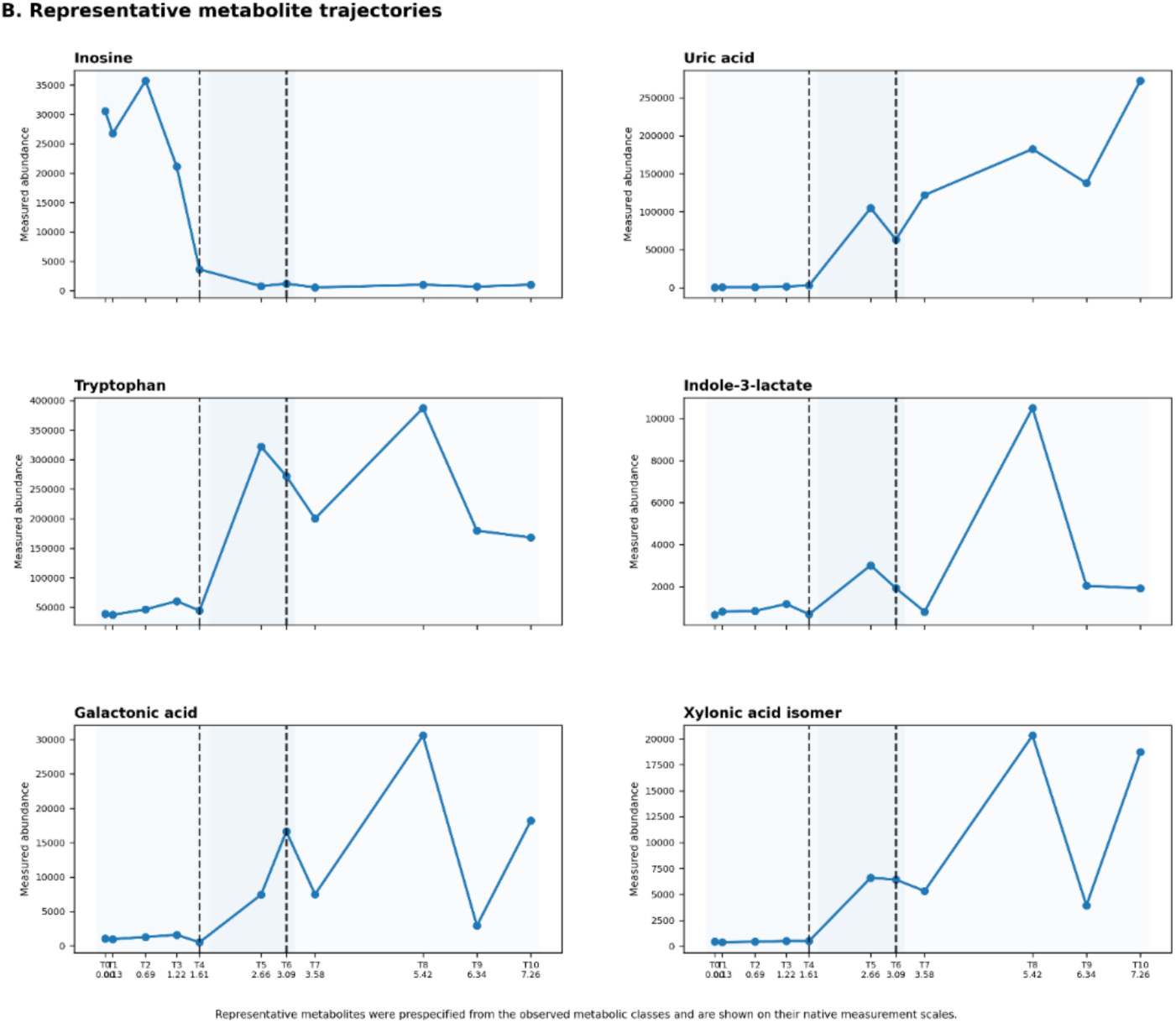
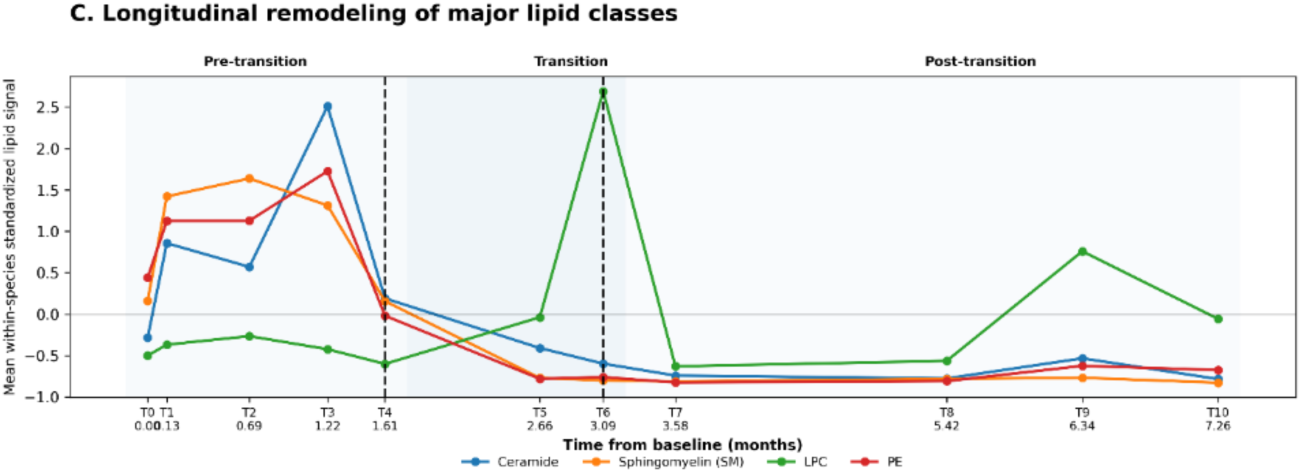
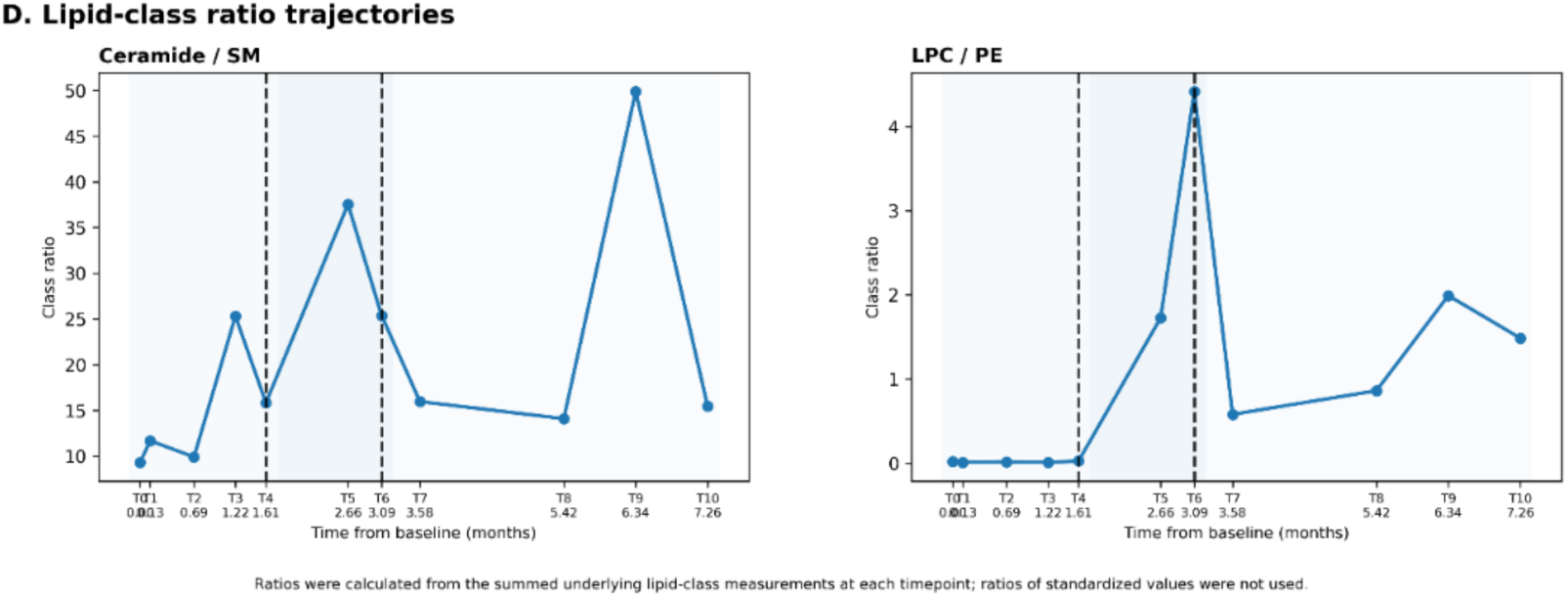
Metabolic transformation of the intestinal environment accompanies the microbial-state transition. (A) Longitudinal remodeling of the stool metabolome. Heatmap showing the 40 metabolites with the greatest longitudinal variability across stool samples T0–T10. Metabolites were selected using a scale-aware variability criterion based solely on the longitudinal metabolomics data and independently of clinical outcome. Selected metabolites were standardized individually across time; therefore, colors represent abundance relative to the longitudinal mean of each metabolite rather than absolute differences in concentration between metabolites. Metabolites were organized to facilitate visualization of biological groups including purine-related, tryptophan/indole-related, sugar-acid/carbohydrate-related, amino-acid-related, and central-carbon metabolites. Samples are displayed chronologically and divided into the same pre-transition (T0–T4), transition (T5–T6), and post-transition (T7–T10) periods defined for the microbial analyses in Figure 2. Light shading and dashed vertical boundaries identify the transition interval. (B) Representative metabolite trajectories. Longitudinal trajectories of six metabolites selected to illustrate prominent changes across major metabolic classes: inosine and uric acid (purine metabolism), tryptophan and indole-3-lactate (tryptophan/indole metabolism), and galactonic acid and xylonic acid isomer (carbohydrate-derived sugar acids). Values are displayed on their native measurement scales rather than as standardized values. The trajectories illustrate distinct temporal patterns, including marked loss of the early inosine signal and emergence or elevation of several metabolites during and after the transition interval. Representative metabolites were selected to illustrate the major metabolic processes evident in the longitudinal dataset and were not selected according to correlation with tumor burden. (C) Longitudinal remodeling of major lipid classes. Class-level trajectories are shown for ceramides, sphingomyelins (SM), lysophosphatidylcholines (LPC), and phosphatidylethanolamines (PE). Individual lipid species were standardized independently across T0–T10 before averaging within each lipid class, preventing lipid species with high absolute signal intensities from disproportionately determining the class trajectory. The resulting values therefore represent coordinated longitudinal changes among lipid species within each class rather than absolute class concentrations. Pronounced lipid remodeling occurred around the microbial/metabolic transition interval, with different lipid classes exhibiting distinct temporal responses. (D) Longitudinal lipid-class signal ratios. Ceramide/sphingomyelin (Cer/SM) and lysophosphatidylcholine/phosphatidylethanolamine (LPC/PE) ratios were calculated at each timepoint from the summed internal-standard-normalized relative intensities of individual lipid species assigned to the corresponding classes. Ratios were calculated before within-species standardization. Because the measurements represent relative analytical responses rather than calibrated molar concentrations, the ratios describe changes in relative lipid-class signals and should not be interpreted as absolute molar ratios or direct biochemical conversion rates. For all panels, time is expressed as months relative to the first longitudinal stool collection (T0 = month 0) to preserve participant confidentiality. The pre-transition, transition, and post-transition designations are identical to those used in Figure 2, allowing direct temporal comparison of microbial and chemical changes. Together, the data demonstrate that the intestinal microbial transition was accompanied by broad remodeling of the stool metabolome and lipidome, including prominent changes in purine-, tryptophan/indole-, carbohydrate-related metabolites and sphingolipid-and phospholipid-associated lipid classes.

**Figure 4.**
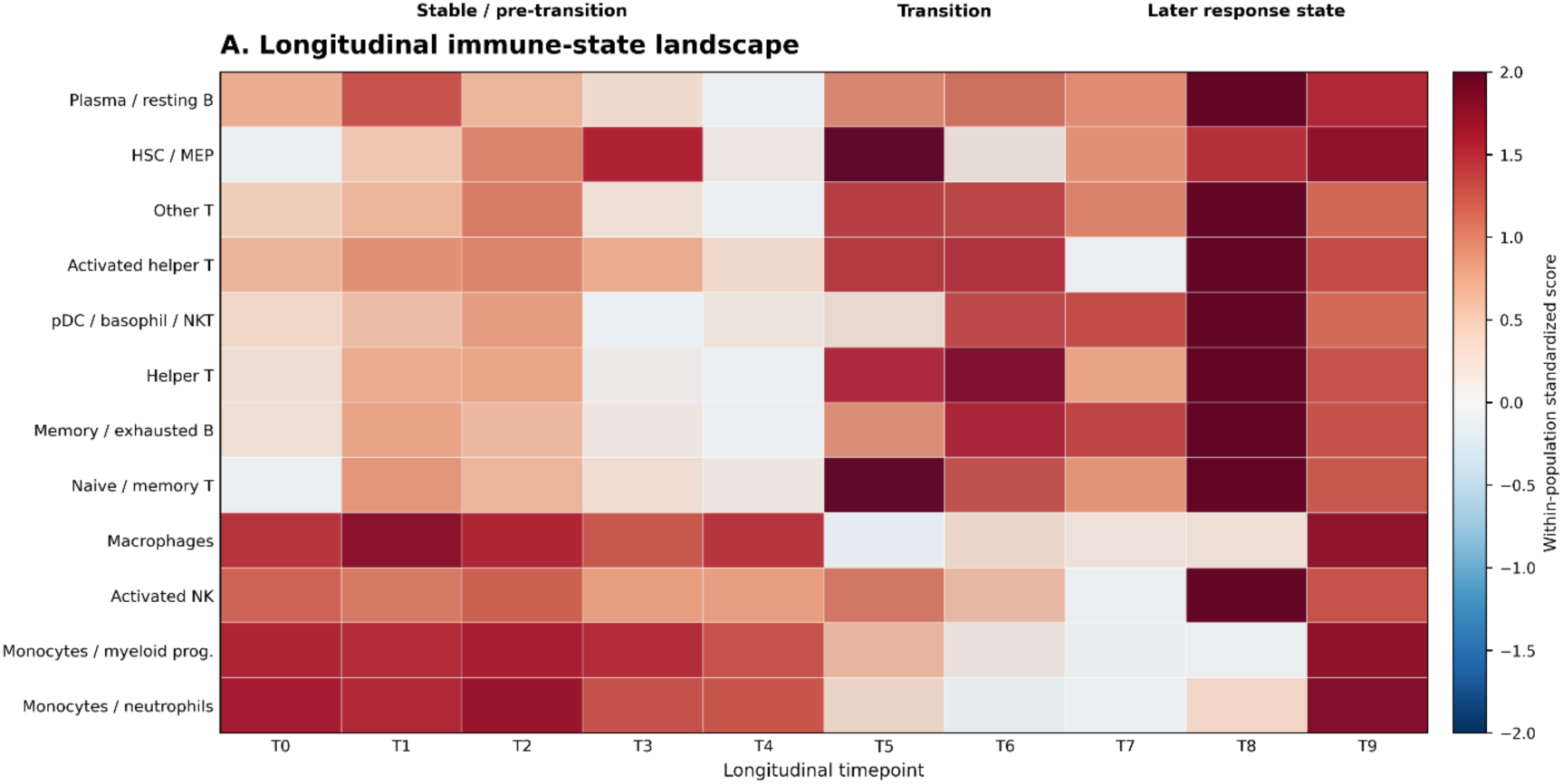
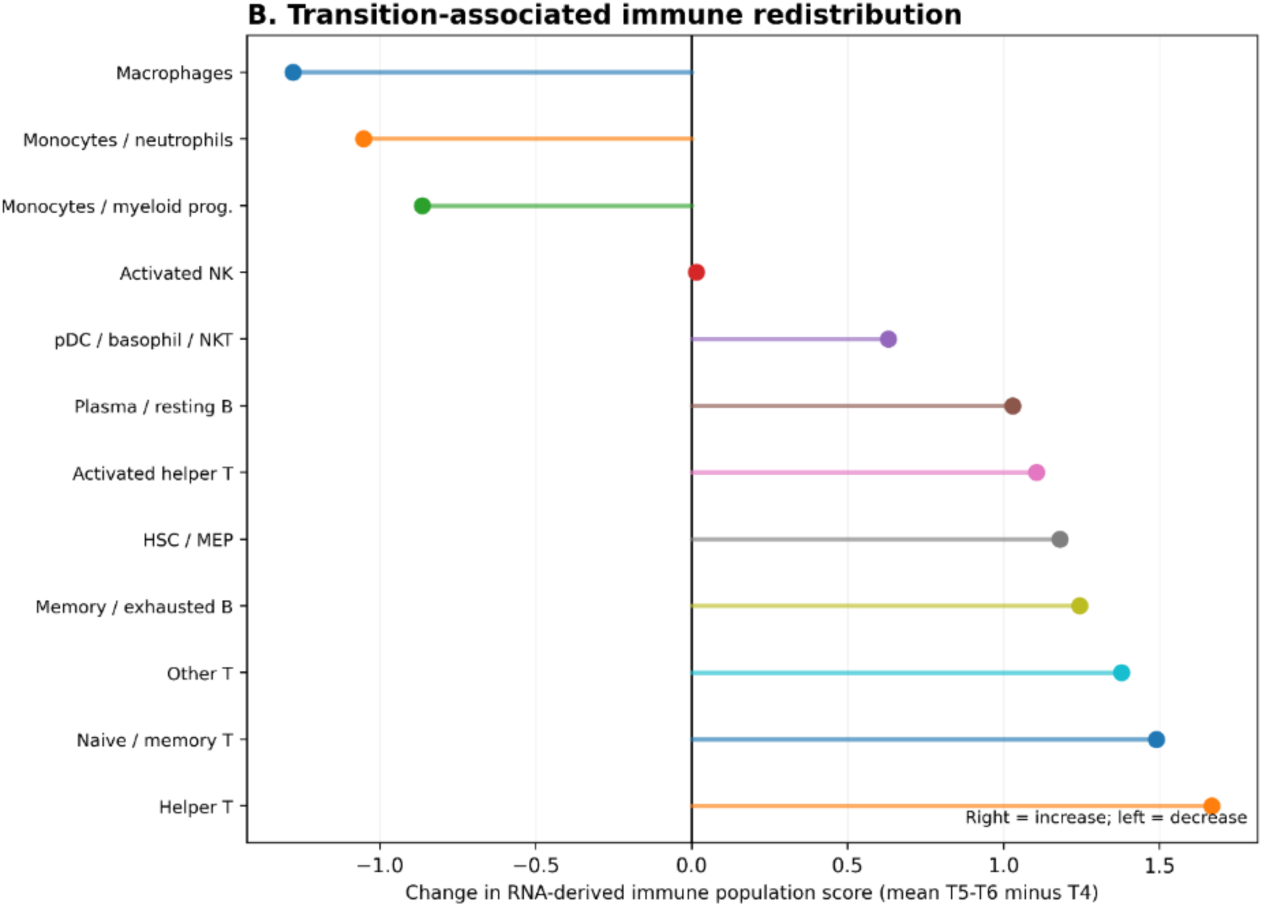
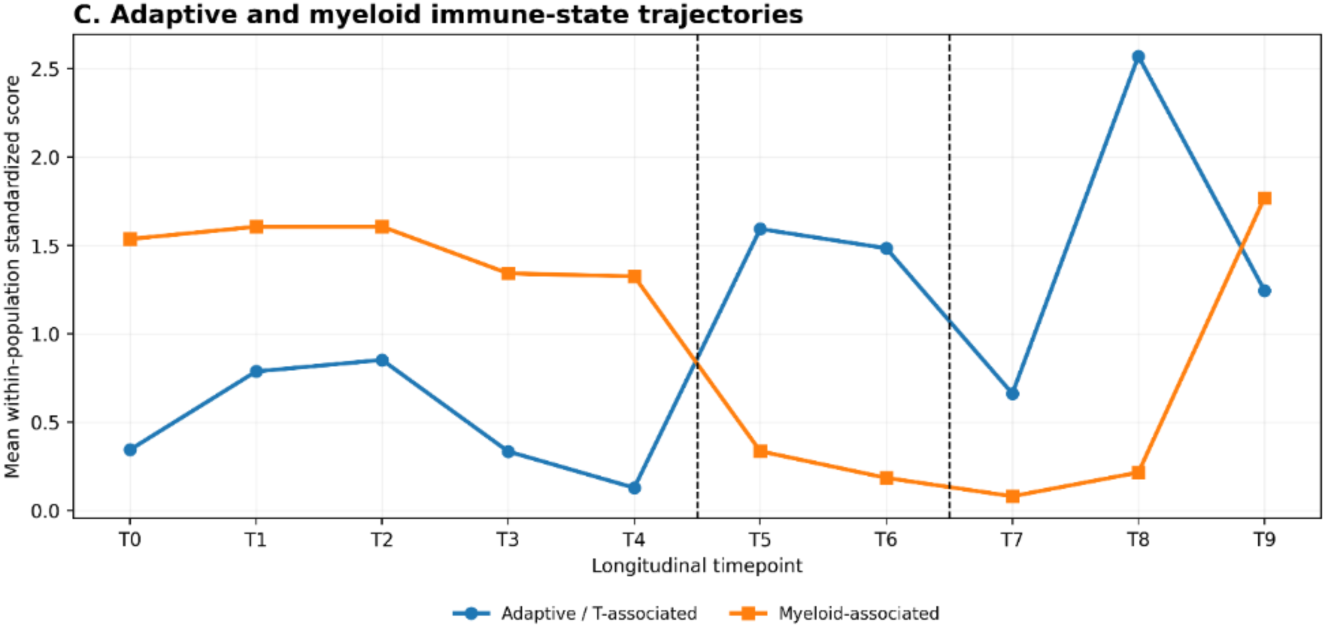
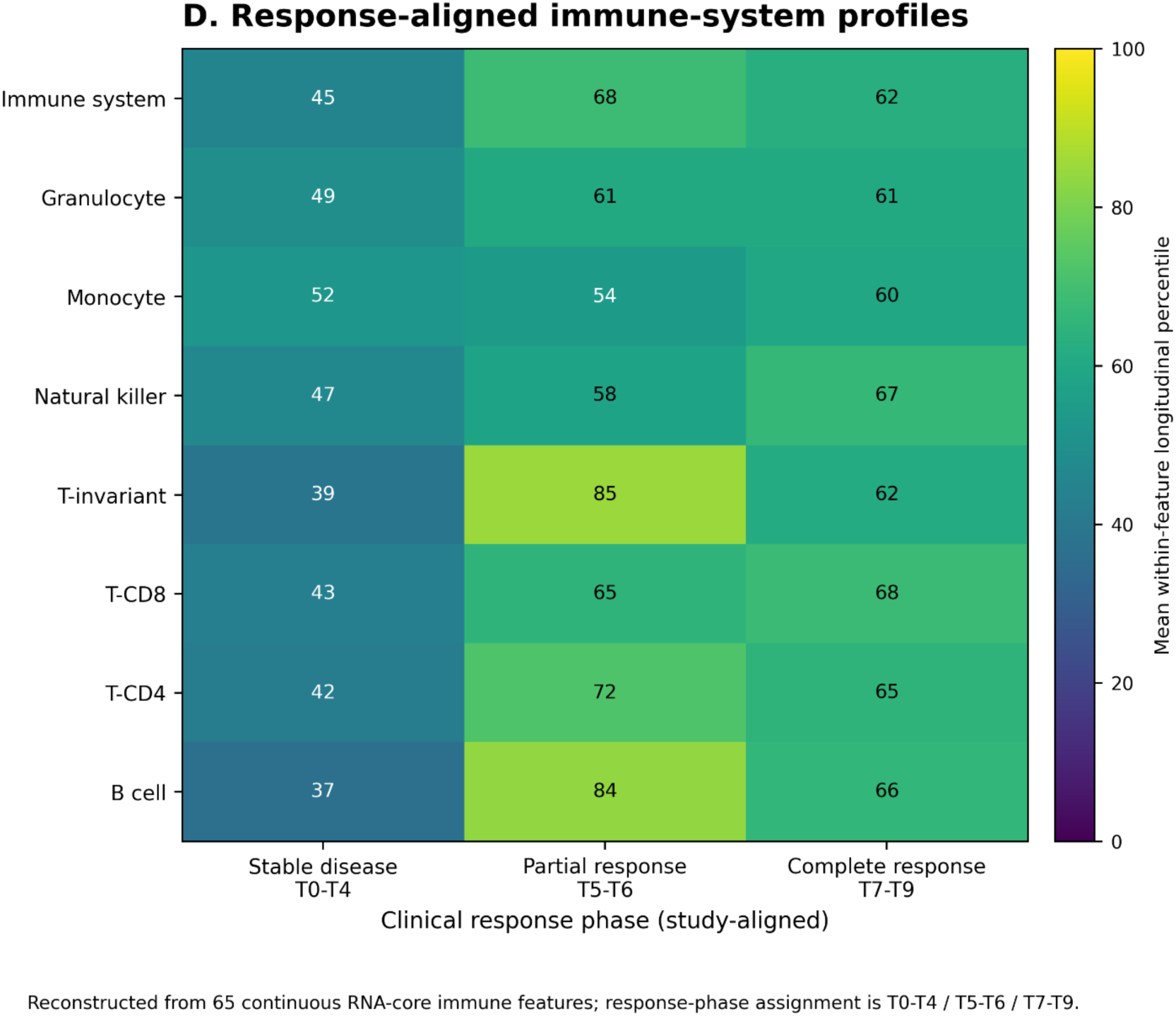
**Longitudinal remodeling of the intestinal host immune landscape during immune checkpoint therapy (A) Longitudinal immune-population trajectories**. Heatmap shows RNA-derived scores for 12 immune population/signature groups across T0–T9. Each population is displayed across the complete longitudinal series to preserve temporal structure rather than averaging measurements into response phases. Colors represent standardized longitudinal activity within each immune signature. The major T5–T6 transition is characterized by increased T-cell-and B-cell-associated signatures together with pronounced reductions in macrophage-and monocyte-associated signatures. Later samples retain features of the altered immune state while undergoing additional compartment-specific remodeling. Immune measurements represent stool host-RNA-derived population/state signatures and not direct immune-cell counts. (B) Transient-associated immune redistribution. Diverging lollipop plot shows the change in each of the 12 immune population scores from the final pre-transition sample (T4) to the mean of T5–T6. Positive values indicate increased RNA-derived signature activity and negative values indicate decreased activity. Helper T, naïve/memory T, other T, activated helper T, and several B-cell-associated signatures increase during the transition, whereas macrophage, monocyte/neutrophil, and monocyte/myeloid-progenitor signatures decline markedly. The zero reference line denotes no transition-associated change. (C) Longitudinal adaptive/T-associated and myeloid-associated immune-state trajectories. The adaptive/T-associated composite is the unweighted mean of activated helper T, helper T, naïve/memory T, and other T scores; the myeloid-associated composite is the unweighted mean of macrophage, monocyte/myeloid-progenitor, and monocyte/neutrophil scores. The two trajectories demonstrate reciprocal reorganization around T5–T6, with increasing adaptive/T-associated activity and decreasing myeloid-associated activity, followed by additional later-state remodeling. The composites are descriptive summaries of predefined constituent populations and were not optimized against clinical response. (D) Response-aligned immune-system profiles. Heatmap summarizes eight host immune-system models reconstructed from 65 longitudinal RNA-derived immune features: overall immune, granulocyte, monocyte, natural-killer, T-invariant, CD8 T-cell, CD4 T-cell, and B-cell systems. Values represent mean within-feature longitudinal percentile scores aggregated within each model across early (T0–T4), transition (T5–T6), and later (T7–T9) intervals. Higher values indicate relatively greater immune-signature activity within the corresponding model and longitudinal series, rather than absolute immune-cell abundance. This response-aligned representation is descriptive; the original continuous immune measurements were retained for quantitative integration in Figure 5. The three intervals represent repeated observations from one individual, not independent clinical-response groups. For all panels, immune values are derived from stool host RNA and represent transcriptionally inferred immune population or state signatures rather than direct cell counts. No T10 immune measurement was available or imputed. The 12-population analysis and 65-feature/eight-model analysis provide complementary levels of resolution of the same longitudinal host immune reorganization.

**Figure 5.**
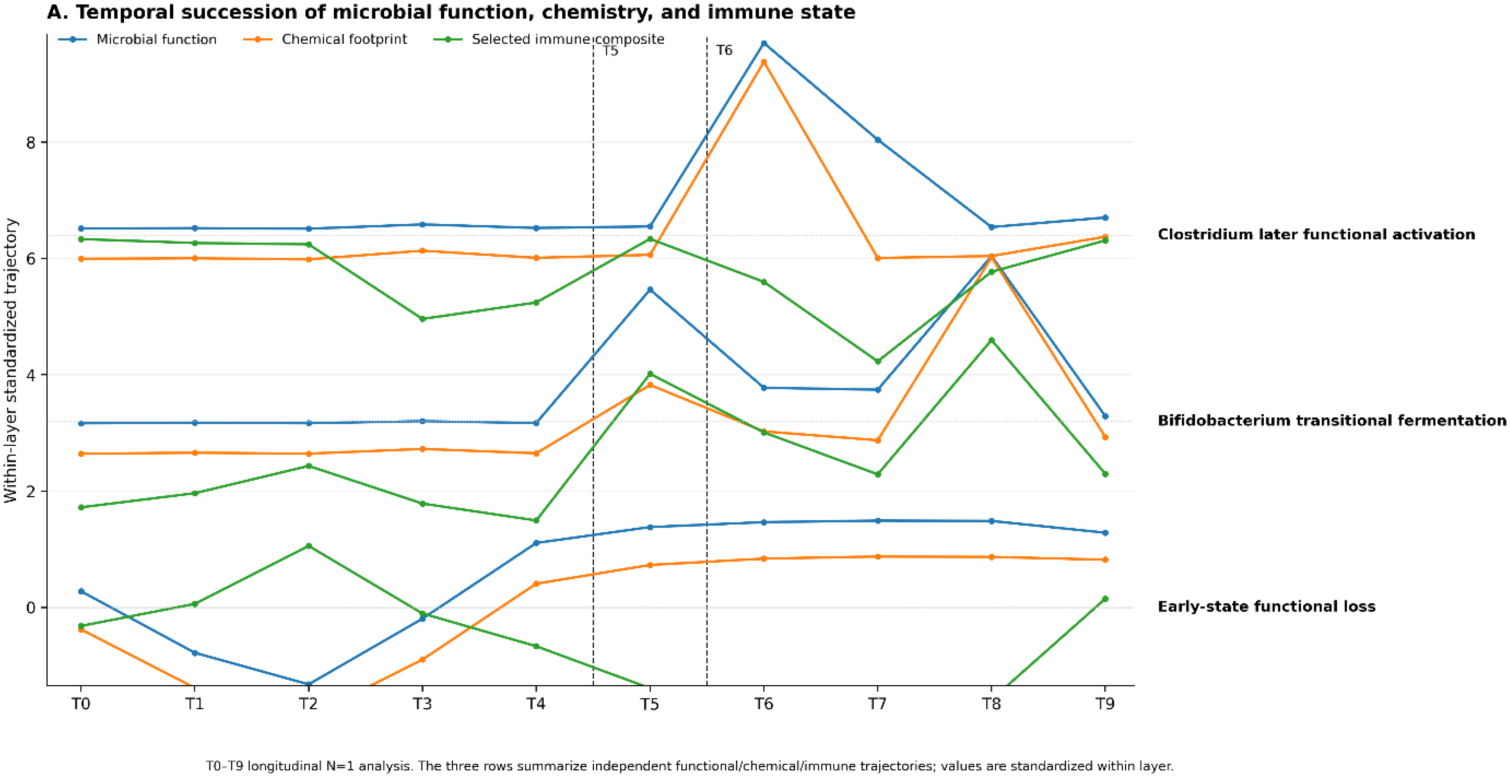
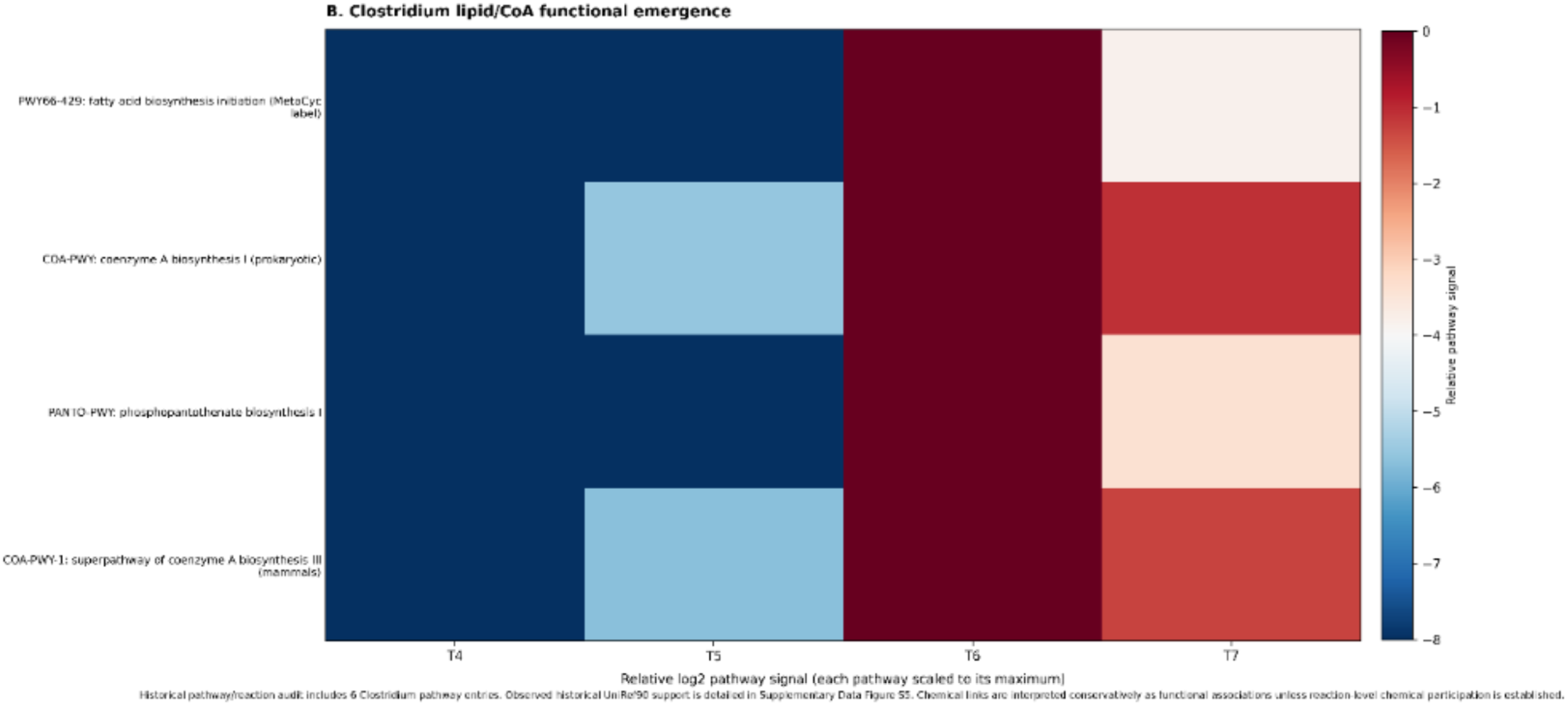
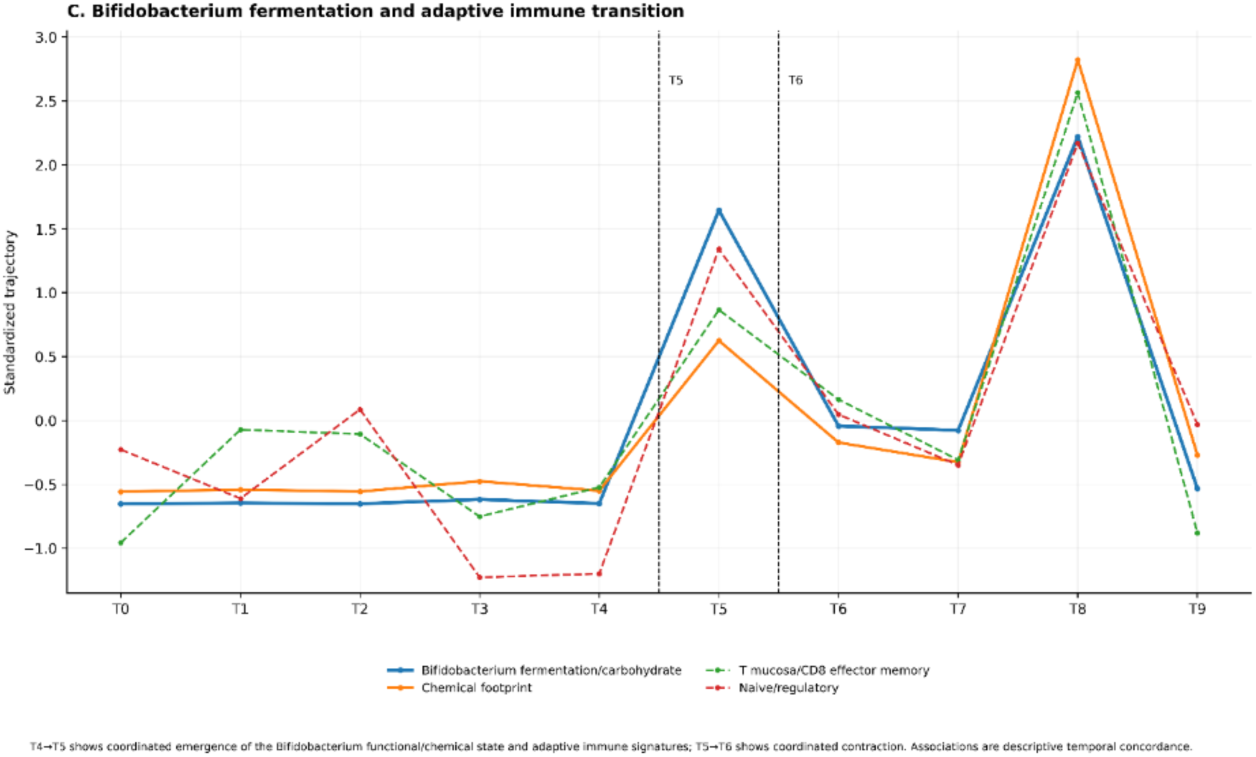
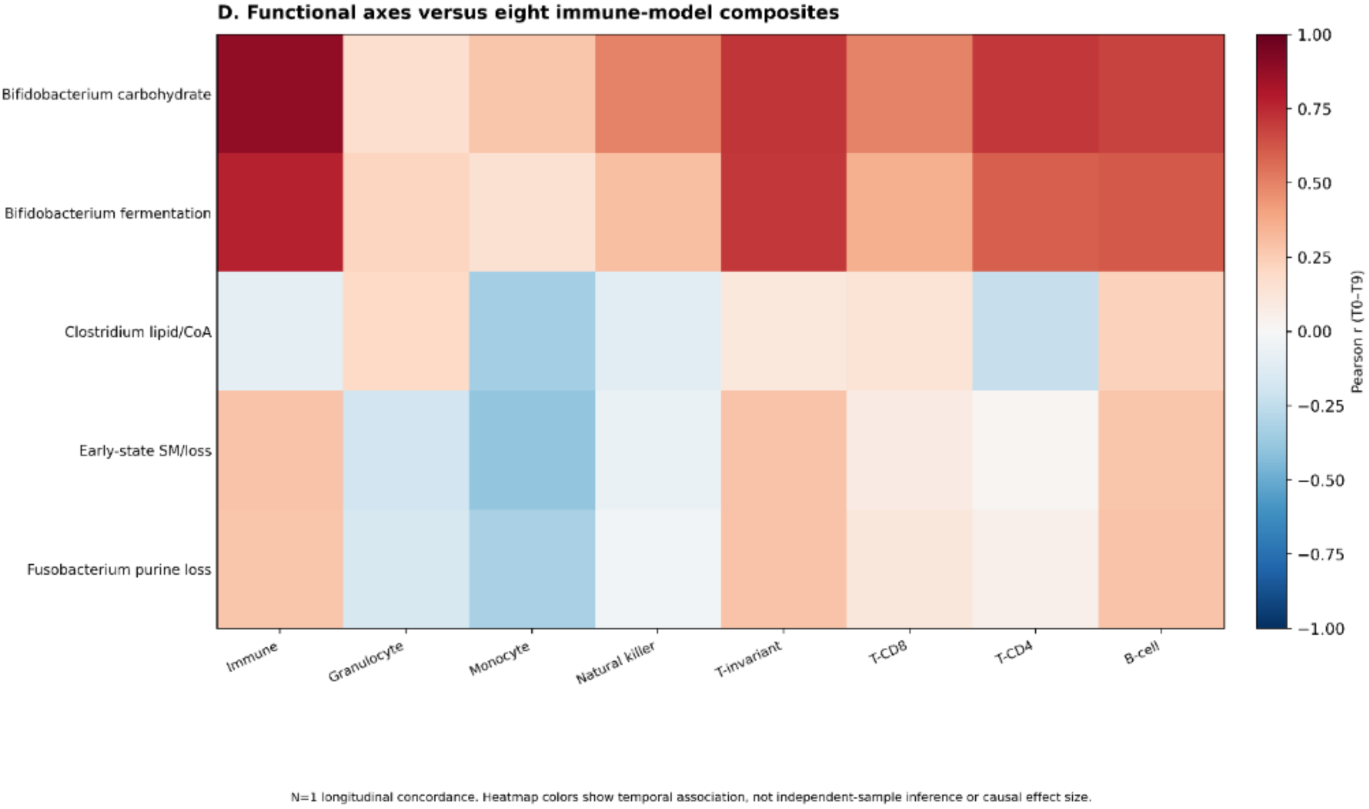
Temporal microbial functional succession, chemical remodeling, and immune-state reorganization. (A) Temporal succession of microbial functional, chemical, and host immune states. Standardized longitudinal trajectories (T0–T9) summarize three nonredundant biological states identified through taxon-resolved metatranscriptomic, metabolomic/lipidomic, and immune integration. The early-state loss axis captures declining *Bacteroides/Fusobacterium*-associated microbial functions together with their associated chemical footprint and selected immune signatures. The *Bifidobacterium* axis represents carbohydrate/fermentation-associated functional activity, its measured chemical footprint, and the corresponding adaptive immune signature. The *Clostridium* axis represents lipid/coenzyme A (CoA)-associated functional activity, fatty-acid/lysophospholipid remodeling, and selected immune signatures. Vertical markers emphasize the T4→T5 and T5→T6 transition windows. The trajectories reveal temporally distinct phases, with early-state functional loss and a *Bifidobacterium*-centered transition at T5 followed by marked *Clostridium* functional activation at T6. (B) Emergence of Clostridium-associated lipid/coenzyme A functional activity. Heatmap shows longitudinal changes in selected taxon-stratified MetaCyc lipid/coenzyme A-associated pathway signals across T4–T7, expressed relative to each pathway’s maximum observed signal on a log2 scale. Historical reconstruction of the original HUMAnN pathway-to-reaction-to-UniRef90 mappings identified patient-observed, *Clostridium*-associated gene-family evidence supporting these functional assignments (Supplementary Table S15). The associated fatty-acid and lysophospholipid measurements are documented in the pathway–chemical audit (Supplementary Table S16). These findings support functional and temporal associations but do not establish metabolic flux or direct bacterial production of the measured lipid species. (C) *Bifidobacterium* fermentation-associated activity coincides with a T5-centered chemical and adaptive immune transition. Longitudinal trajectories show *Bifidobacterium*-associated pyruvate fermentation/carbohydrate activity together with measured fermentation-associated chemicals, including lactate and indole-3-lactate, and representative RNA-derived adaptive immune signatures. The microbial functional, chemical, and immune layers rise sharply from T4 to T5 and subsequently decline toward T6, defining a temporally coherent transitional state. Representative immune features were selected from the predefined 65-feature core immune analysis on the basis of the immune-integration criteria and include T-helper, memory/regulatory, mucosal/effector, and related adaptive signatures. Historical MetaCyc reaction-to-UniRef90 mapping provides molecular support for the microbial functional program; the association with indole-3-lactate is interpreted as temporal chemical concordance rather than direct production by the displayed fermentation pathway. (D) Distinct microbial functional states associate with different intestinal immune compartments. Heatmap summarizes longitudinal associations between the five predefined, nonredundant microbial functional/chemical axes and eight core immune-system composite trajectories derived from the comprehensive RNA-core immune analysis. Colors indicate longitudinal Pearson correlation coefficients across T0–T9. The *Bifidobacterium*carbohydrate/fermentation axes show broad positive relationships with T-cell, T-invariant, and B-cell-associated immune programs, whereas *Clostridium* lipid/CoA activation exhibits a more selective immune relationship. Loss of the early *Bacteroides/Fusobacterium* functional state is associated predominantly with remodeling of monocyte/myeloid and related innate immune signatures. Immune measurements represent stool host-RNA-derived population/signature scores and should not be interpreted as direct cell counts. Correlations describe temporal concordance within this longitudinal N-of-1 study and do not establish causality or population-level association.

Representative metabolites displayed marked, unstandardized trajectory shifts across the transition interval (Fig. 3B; Supplementary Table S6). In purine metabolism, inosine was enriched early and declined sharply during transition, whereas uric acid increased substantially. Tryptophan and the microbial metabolite indole-3-lactate increased during the transition, aligning with the timing of microbial restructuring (Fig. 2). Carbohydrate-derived metabolites, including galactonic acid and xylonic acid isomer, showed similarly abrupt alterations, demonstrating that biochemical remodeling extended across both microbial and central host-microbial intermediary pathways.

Lipidomic profiling revealed parallel remodeling across major stool lipid classes (Fig. 3C,D). To evaluate class-level dynamics without bias from high-abundance species, individual lipid species were standardized before averaging into four primary classes: ceramides, sphingomyelins (SM), lysophosphatidylcholines (LPC), and phosphatidylethanolamines (PE). Ceramides and sphingomyelins shifted prominently around the transition interval, alongside distinct changes in LPC and PE (Fig. 3C). Analysis of individual constituent species confirmed that these shifts reflected class-wide alterations rather than isolated single-lipid driven effects (Supplementary Fig. S6; Supplementary Tables S7 and S9).

To assess changes in the quantitative balance among related lipid families, we evaluated class ratios using summed, non-z-standardized internal-standard-normalized intensities (Fig. 3D; Supplementary Table S10). Both Ceramide/SM and LPC/PE ratios exhibited marked longitudinal shifts across the treatment course, indicating changes in the relative distribution of measured lipid classes.

Together, these findings demonstrate that the longitudinal microbial transition extended directly into the chemical lumen of the gut. The timing of this metabolic and lipidomic transition closely paralleled the microbial shift (Fig. 2) and coincided with the emergence of sustained clinical response (Fig. 1). These coordinated multi-omic trajectories provided the rationale for subsequent functional metatranscriptomic analyses linking specific microbial metabolic pathways to these observed chemical and host immune changes.

### Longitudinal host-RNA profiling reveals coordinated but nonuniform remodeling of the intestinal immune state

Longitudinal stool host-RNA profiling across T0-T9 revealed marked but nonuniform changes in immune-associated transcriptional signatures (Fig. 4). The transition was characterized by reciprocal changes in adaptive/lymphoid-associated and myeloid-associated programs rather than a nonspecific increase in all immune signals. Among 12 RNA-derived immune population/state signatures, the early period (T0-T4) showed relatively prominent macrophage/monocyte-associated signals. At T5-T6, helper T, activated helper T, naïve/memory T, and several B-cell-associated signatures increased, whereas macrophage and monocyte-associated signatures declined (Fig. 4A). Comparison of T4 with the mean of T5-T6 summarized the immediate redistribution (Fig. 4B). Descriptive adaptive/T-associated and myeloid-associated composites showed opposing longitudinal trajectories (Fig. 4C). Individual T4, T5, T6, and T8 profiles highlighted continuing changes after the initial transition (Supplementary Fig. S7). These RNA signatures should not be interpreted as direct cell counts. The complete 12-signature longitudinal matrix is provided in Supplementary Table S11.

The expanded 65-feature, eight-model immune analysis (overall immune, granulocyte, monocyte, natural-killer, T-invariant, CD8 T, CD4 T, and B-cell systems) showed complementary response-aligned patterns (Fig. 4D; Supplementary Fig. S8). T-and B-associated models generally showed greater relative signal in later response-aligned states, whereas monocyte and other innate programs exhibited distinct, nonuniform behavior. These are descriptive summaries of repeated measurements from one individual, not independent clinical-response groups. This host-RNA reorganization overlapped the microbial and chemical transitions (Figs. 2 and 3), motivating integration of the continuous immune trajectories with independently selected microbial functional and chemical axes. Continuous 65-feature measurements and response-aligned eight-model percentile summaries are provided separately in Supplementary Tables S12 and S13, respectively.

### Temporal microbial function, chemical remodeling, and immune-state reorganization

To investigate the functional basis of the intestinal ecosystem transition, we integrated genus-harmonized 16S abundance, taxon-stratified metatranscriptomic activity, MetaCyc pathways, measured metabolites and lipids, and longitudinal host-RNA immune signatures.

Microbial taxa and functional pathways were prioritized independently of the chemical and immune measurements, and redundant pathway-chemical associations were consolidated into nonredundant functional axes. This approach identified temporally distinct microbial-metabolic-immune states rather than a single coordinated transition (Fig. 5A; Supplementary Data Fig. S9). The full functional-axis consolidation and pathway-membership audit is provided in Supplementary Table S14.

The earliest component involved loss of the transcriptionally active *Bacteroides/Fusobacterium*-associated microbial state, accompanied by purine-and sphingomyelin-associated chemical remodeling. These loss-associated functional axes were prominent before the transition and declined as the intestinal ecosystem reorganized (Supplementary Data Fig. S9B,C). Their disappearance coincided with sustained changes in host monocyte-, myeloid-, neutrophil-, and NK-associated RNA signatures, suggesting that loss of the early microbial functional environment was associated with reorganization of the intestinal innate immune state.

A distinct *Bifidobacterium*-associated functional state emerged at T5. Increased carbohydrate and fermentation-associated transcriptional activity coincided with changes in measured fermentation-related chemicals, including lactate and indole-3-lactate, and pronounced remodeling of adaptive immune signatures (Fig. 5A,C). The strongest immune relationships involved T-helper-17/γδ-associated memory, mucosal/CD8 effector-memory, terminal-memory/regulatory, and naïve/regulatory T-cell signatures. These relationships showed strong longitudinal and first-difference concordance, with representative correlation coefficients generally exceeding 0.8. Importantly, microbial functional activity, its chemical footprint, and the associated adaptive immune signatures increased together from T4 to T5 and declined from the T5 peak toward T6, with additional later variation including a second rise near T8. This pattern identifies T5 as a transient microbial–metabolic–adaptive immune state rather than a sustained increase in *Bifidobacterium* activity. Targeted pathway–chemical evidence and functional-axis/immune-feature associations are documented in Supplementary Tables S16 and S17.

The subsequent T5→T6 interval was characterized by marked emergence of a different functional program associated with Clostridium (Fig. 5A,B). Taxon-stratified MetaCyc profiles demonstrated activation of lipid and coenzyme A (CoA)-associated pathways, including fatty-acid biosynthesis initiation, pantothenate metabolism, and CoA biosynthesis. These pathways were minimally detected at T4, became detectable at T5, and showed pronounced activity at T6. Historical reconstruction using the original HUMAnN v3.9, UniRef90 2019_01, and MetaCyc v24 mapping resources confirmed that the prioritized microbial functions were supported by taxon-specific, reaction-associated UniRef90 families observed in the patient’s metatranscriptomic data. The *Clostridium* functional transition coincided with substantial remodeling of measured fatty-acid and lysophosphatidylcholine species. Although these observations provide molecular support for microbial functional activation and strong temporal association with the lipidomic changes, they do not establish direct bacterial production of the measured lipids. Reaction-associated molecular verification and observed UniRef90 evidence are detailed in Supplementary Table S15.

The *Clostridium*-associated state also exhibited an immune pattern distinct from the preceding *Bifidobacterium* transition. Whereas the T5 fermentation-associated state tracked broad adaptive immune remodeling, later *Clostridium* lipid/CoA activity was positively associated with a granulocyte-progenitor signature and inversely associated with selected monocyte-associated signatures (Fig. 5A,D). Across the eight immune-model composites, the two *Bifidobacterium* axes showed broadly positive relationships with overall immune, T-invariant, CD4, CD8, and B-cell-associated programs. In contrast, the *Clostridium* and early microbial-loss axes exhibited more selective and heterogeneous relationships, particularly involving monocyte and other innate immune compartments. These differences indicate that the microbial functional transition was accompanied by redistribution among immune programs rather than uniform immune activation.

The molecular and chemical audit further supported this interpretation (Supplementary Data Fig. S9A-D). Historical pathway-to-reaction-to-UniRef90 reconstruction identified patient-observed molecular evidence for the prioritized microbial functional programs, while chemical membership analysis distinguished measured metabolites and lipids associated with each axis. Importantly, none of the retained pathway-chemical relationships was classified as directly verified at the reaction-chemical level. Consequently, the chemical footprints were interpreted as functional associations or ecosystem-state markers rather than confirmed microbial products.

The complete 65-feature immune evidence analysis similarly demonstrated that the associations were concentrated within particular immune signatures rather than distributed uniformly across the host immune landscape (Supplementary Data Fig. S9E). The T4→T5 and T5→T6 stage-specific changes and supporting longitudinal trajectories are provided in Supplementary Table S18.

Together, these analyses resolve the intestinal transition into three overlapping but temporally distinguishable components: loss of an early microbial functional state, a T5-centered *Bifidobacterium* fermentation/adaptive-immune transition, and a subsequent T6-centered *Clostridium* lipid/CoA-associated state. Their distinct chemical footprints and immune relationships suggest that treatment-associated intestinal remodeling involved microbial functional succession rather than simple enrichment of a favorable bacterial taxon. Although these changes occurred during sustained clinical tumor regression, the single-subject observational design does not establish causality. Instead, the convergence of microbial transcriptional activity, molecularly supported functional pathways, measured chemistry, and host immune signatures identifies specific microbial–metabolic–immune relationships for testing in larger longitudinal ICI cohorts.

## Discussion

Dense longitudinal multi-omics integration of a patient with MSI-H/dMMR colorectal cancer metastatic to the liver revealed temporally distinct microbial, chemical, and intestinal immune-state changes during response to nivolumab plus ipilimumab therapy. Rather than enrichment of a single response-associated organism, the principal finding was microbial functional succession accompanied by distinct chemical and host immune signatures. Loss of an early *Bacteroides/Fusobacterium*-associated functional state was followed by a transient *Bifidobacterium* carbohydrate/fermentation state centered at T5 and subsequent emergence of a *Clostridium* lipid/coenzyme A (CoA)-associated state at T6. Importantly, microbial abundance and transcriptional activity were frequently discordant, suggesting that dynamic functional states may provide a more informative representation of microbiome remodeling during ICI therapy than taxonomic composition alone.

The temporal separation of T5 and T6 was particularly informative. At T5, *Bifidobacterium*-associated carbohydrate and fermentation activity increased together with fermentation-associated metabolites and broad adaptive immune remodeling involving T-helper-, memory/regulatory-, mucosal/effector-, and B-cell-associated transcriptional signatures. These signals subsequently declined toward T6. In contrast, the principal *Clostridium* transition occurred between T5 and T6, when taxon-attributed RNA, lipid/CoA-associated pathways, and a fatty-acid/lysophosphatidylcholine (LPC) chemical footprint increased markedly. This later state was associated with a more selective immune configuration, including increased granulocyte-progenitor-associated activity and decreased monocyte-and selected NK/T-cell-associated signatures. Historical reconstruction of the original HUMAnN pathway-to-reaction-to-UniRef90 relationships provided patient-specific molecular support for the prioritized microbial functional programs, although these observations establish neither metabolic flux nor direct bacterial production of the measured chemicals. Together, these findings distinguish a transient fermentation-associated adaptive immune state from a subsequent lipid/CoA-associated state with a different pattern of host immune remodeling.

Experimental studies provide biological context for these distinct immune signatures. Commensal *Bifidobacterium* has been shown to enhance dendritic-cell function and CD8 T-cell priming during PD-L1 blockade, while *B. pseudolongum*-derived inosine can modulate checkpoint efficacy through context-dependent T-cell signaling [10, 37]. Selected commensal Clostridia have been shown to promote intestinal regulatory T-cell differentiation, partly through microbial fermentation products such as butyrate, whereas defined commensal consortia can also enhance intestinal CD8 T-cell responses and checkpoint efficacy [38–40]. These findings demonstrate that microbial metabolic activity can influence different immune programs, although the specific pathways responsible depend on the organism and biological context. The *Clostridium* lipid/CoA-associated state identified here cannot be equated with butyrate production or regulatory T-cell induction, and the observed immune signatures do not establish that these published mechanisms operated in our patient.

Our findings also extend previous studies linking intestinal microbiota to ICI efficacy [3–8]. Cross-cohort investigations have demonstrated limited reproducibility of individual response-associated taxa [12], while longitudinal studies show that microbial composition and function continue to change after treatment initiation [13, 15, 17, 18]. Our study adds a complementary perspective by resolving the temporal relationships among microbial transcriptional activity, measured metabolites and lipids, and host immune signatures within the same individual. The observed abundance-activity discordance suggests one possible explanation for inconsistent taxonomic associations: different microbial communities may perform overlapping metabolic functions, while individual organisms may occupy distinct transcriptional states depending on ecological and treatment context. Thus, microbial functional states may provide a more informative framework for investigating microbiome-ICI interactions than taxonomic composition alone.

An important unresolved question is what drives the observed microbial functional succession during checkpoint blockade. Experimental evidence suggests that ICI treatment can modify intestinal microbial ecology through host-mediated mechanisms. In mouse models, PD-1 blockade has been reported to alter intestinal IgA responses, bacterial IgA binding, microbial composition, and metabolite production [41], while CTLA-4 blockade can induce microbiota-dependent intestinal immune activation [42, 43]. Conversely, microbial metabolites can influence antitumor immunity, including context-dependent effects of *Fusobacterium nucleatum*-associated metabolites on checkpoint efficacy [11, 38]. The pronounced decline in *Fusobacterium*-associated transcriptional activity observed here could reflect reduced abundance, transcriptional suppression, ecological displacement, or host-mediated selection. Diet, concomitant medications, changing tumor burden, and intestinal physiology may also contribute. These possibilities suggest a potentially bidirectional host–microbiome feedback system, but the directionality and causal relationships remain unresolved.

The N-of-1 design is central to this contribution. Although a single patient cannot establish population-level associations, generalizable biomarkers, or causal mechanisms, dense within-individual sampling addresses questions that conventional cross-sectional studies cannot readily resolve. By comparing each measurement with the patient’s preceding biological state, this approach reduces the influence of between-person heterogeneity and allows transient or sequential changes to be distinguished from sustained responses. In the present study, this resolution was essential for identifying abundance-activity discordance, the T5-centered

*Bifidobacterium* transition, and subsequent T6-centered *Clostridium* functional emergence.

Previous longitudinal personal-omics and microbiome studies similarly demonstrate the value of repeated within-individual measurements for characterizing dynamic molecular states [15, 17–19]. Thus, the principal value of this study lies in generating temporally specific biological hypotheses that can subsequently be evaluated across patients.

These observations generate three related hypotheses for future investigation. First, favorable ICI responses may be accompanied by recurrent microbial functional-state transitions even when the taxa responsible differ among individuals. Second, transient fermentation-associated activity may coincide with an earlier phase of adaptive immune remodeling, whereas subsequent lipid/cofactor-associated activity may characterize a distinct phase of intestinal ecosystem reorganization. Third, disappearance of early microbial functional programs may be as informative as emergence of new programs, particularly in relation to innate and myeloid immune-state changes. Testing these hypotheses in larger longitudinal cohorts will help determine whether patients converge on shared functional and immunometabolic trajectories or instead exhibit highly individualized patterns. Such studies may also establish whether longitudinal microbial functional, chemical, and host immune measurements provide information beyond baseline taxonomic composition and eventually support functional biomarkers or personalized monitoring strategies. Establishing causal mechanisms and potential therapeutic applications will require experimental validation of candidate microbial pathways and molecules.

Several limitations warrant consideration. The observational single-subject design does not establish that microbial or metabolic changes caused tumor regression or immune remodeling. Importantly, the MSI-H/dMMR tumor phenotype provides an established biological basis for sensitivity to checkpoint blockade, and the observed intestinal changes cannot be considered independent determinants of therapeutic response. Treatment effects, changing tumor burden, diet, concomitant medications, intestinal physiology, and reciprocal host– microbiome interactions may all have contributed to the observed trajectories. Furthermore, stool host-RNA measurements represent immune-associated transcriptional signatures rather than direct cell counts or systemic immune measurements. Genus-level taxonomic assignments do not resolve strain-specific functions, and pathway-to-UniRef90 reconstruction does not directly establish enzymatic activity or metabolic flux. Although lipidomic measurements were internal-standard-normalized by the analytical core, pathway–lipid associations do not establish direct bacterial production of individual lipid species. Experimental studies incorporating targeted metabolite quantification, stable-isotope tracing, microbial culture, metaproteomics, and functional perturbation will be needed to evaluate the proposed mechanisms.

In summary, dense longitudinal multi-omics resolved a temporally structured succession of microbial functional, chemical, and intestinal immune states during sustained tumor regression. These findings suggest that microbiome-ICI relationships may be individualized at the taxonomic level while potentially converging on shared functional and immunometabolic states. Testing this possibility in larger longitudinal cohorts may help determine whether dynamic microbial functions and personalized within-patient trajectories provide useful information beyond static taxonomic biomarkers.

## Data Availability

All data produced in the present study are available upon reasonable request to the authors.

## Acknowledgments

This study was supported by funds from Jean Perkins Foundation to E.Z. K. M-E. was supported by the Stanford University Medical Scientist Training Program grant T32-GM007365. We would like to acknowledge the UCLA Goodman-Luskin Microbiome Center Microbiome Core for providing microbiome sequencing services. Metabolomics and lipidomics data were acquired and processed at the UC Davis West Coast Metabolomics Center. We thank Mr. Brian Pierce for retrieving and preparing the clinical CT images used in the longitudinal assessment of tumor regression. His assistance was provided through the CTSI/CRI/HDIP program at the University of Southern California (USC), involving the Southern California Clinical and Translational Science Institute (SC CTSI) and Keck Medicine of USC.

## Author contributions

E.Z. conceived and designed the study, developed the research protocol, performed the integrated microbiome, metatranscriptomic, metabolomic, lipidomic, and immune-related data analyses, conducted the multi-omic integration and computational analyses, prepared all figures and tables, and wrote the original manuscript. E.Z. also supervised the overall study and was responsible for the scientific interpretation and integration of the findings.

K.M.-E. performed RNA sequencing, processed the sequencing data, and conducted the initial host transcriptomic analyses, including computational characterization of immune-cell populations and functional signatures.

H.-J.L. provided clinical care and administered the immunotherapy regimen, contributed the patient’s clinical diagnosis, treatment history, and longitudinal clinical assessments, and provided clinical interpretation of the therapeutic response.

F.A. coordinated stool sample collection, processing, and organization, performed genomic DNA extraction for 16S rRNA sequencing, and conducted the initial 16S sequencing data analyses.

All authors reviewed and edited the manuscript, approved the final version, and agreed to be accountable for their respective contributions to the work.

## Notes

### Competing Interest Statement

The authors have declared no competing interest.

### Author Declarations

This study was conducted under the institutional research protocol entitled The Role of Microbiome in Cancer Therapy (Protocol No. OS-16-9), with oversight by the University of Southern California Institutional Review Board (USC IRB No. HS-CG-13-0003). Longitudinal clinical and multi omics data were analyzed using de-identified study identifiers, with calendar dates replaced by relative time from the first stool collection to protect patient confidentiality. Written informed consent was obtained from the participant in accordance with the approved institutional protocol.

