## Supplementary Figures S1-S9 for "Dense Longitudinal Multi-Omics Reveals Microbial–Metabolic–Immune Succession During Immune Checkpoint Therapy"

**Supplementary Figure S1. Longitudinal CT imaging**

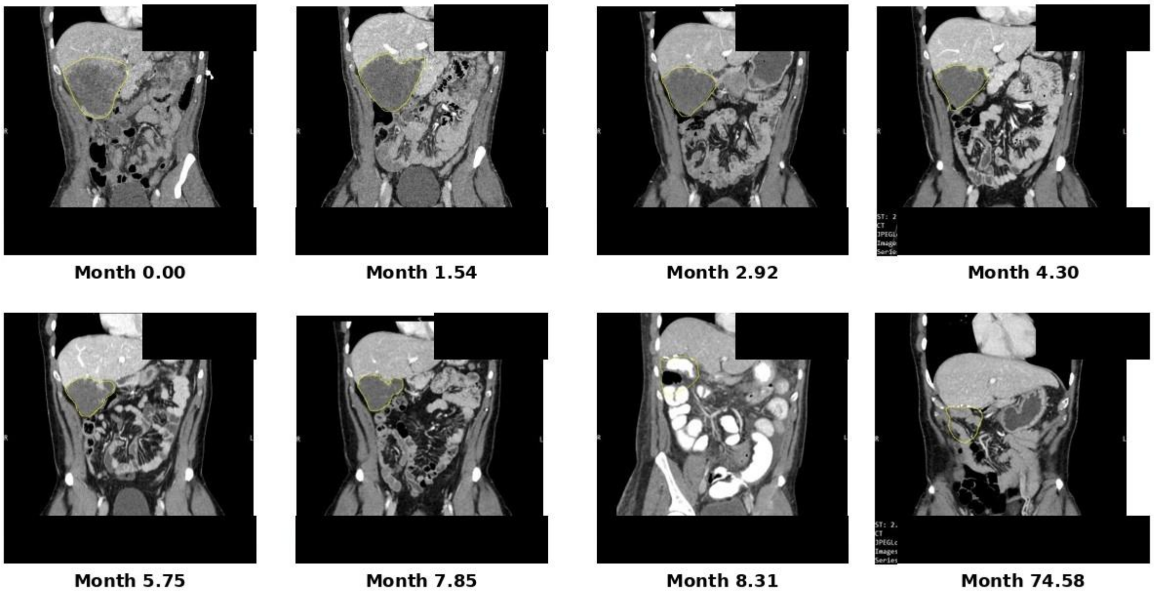


Time is expressed as months from the first CT examination (baseline = month 0).

**Supplementary Figure S1. Longitudinal radiographic response during therapy and long-term follow-up.**

Representative coronal computed tomography (CT) images illustrate the longitudinal evolution of the dominant lesion during treatment and long-term follow-up. To protect participant confidentiality, calendar dates and identifying imaging metadata were removed. Imaging timepoints are expressed as elapsed months relative to the designated imaging baseline (month 0). The relationship between the imaging baseline and the first stool collection (T0) is specified in Figure 1 and the clinical timeline. Serial images are shown at months 0.00, 1.54, 2.92, 4.30, 5.75, 7.85, and 8.31, with long-term follow-up imaging at month 74.58. The lesion is delineated by the yellow outline to facilitate comparison across serial examinations. Progressive reduction in lesion size is evident during therapy, consistent with the relative hepatic lesion-area trajectory presented in Figure 1B. The residual lesion was subsequently surgically resected; therefore, imaging after surgery should not be interpreted as continued spontaneous radiographic regression. The month 74.58 image documents long-term radiographic follow-up.

**Supplementary Figure S2. Expanded longitudinal profiling of bacterial community composition and transcriptional activity.**

**Supplementary Figure S2A. Expanded 16S genus-level community composition**

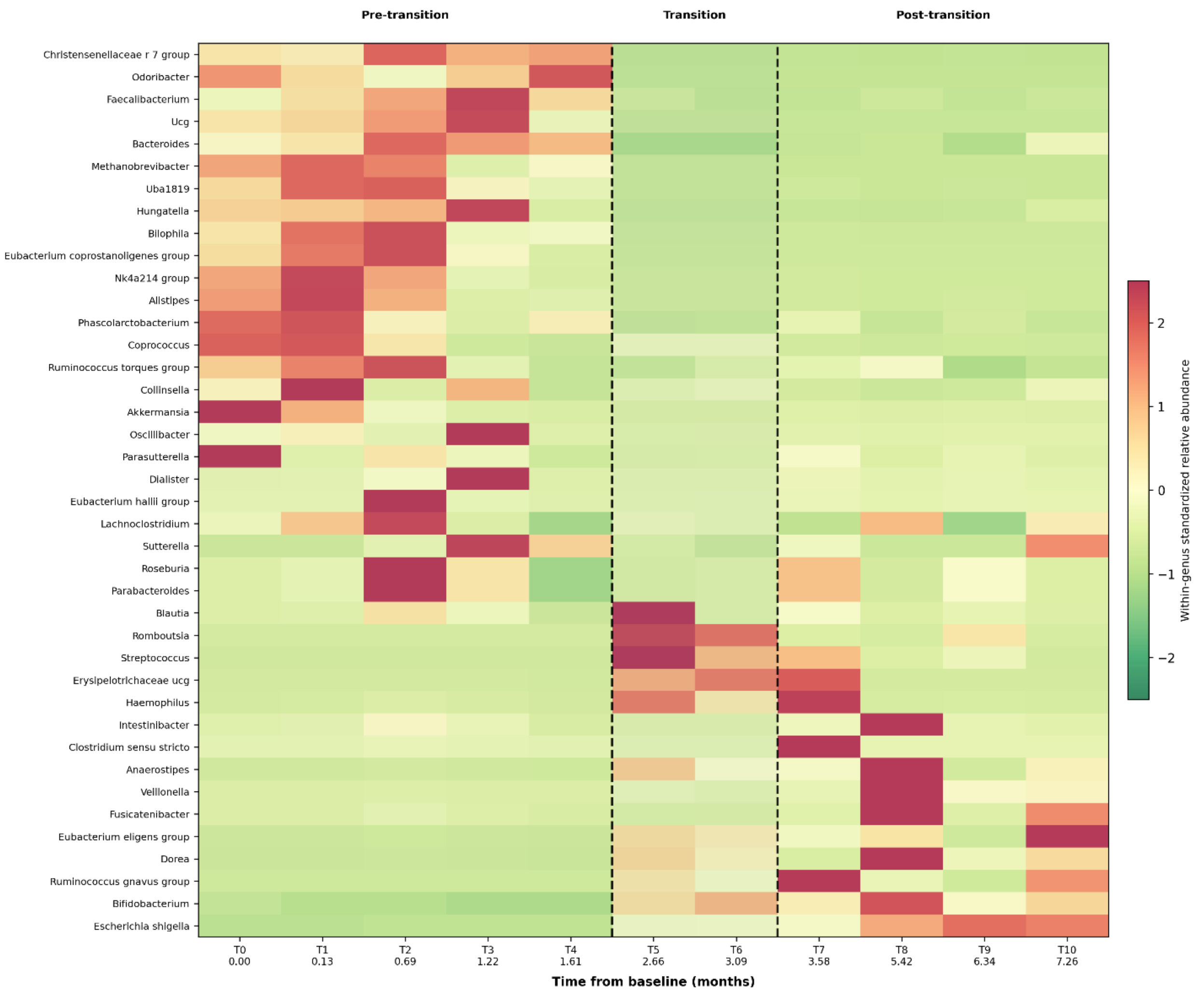


**(A) Expanded 16S genus-level community heatmap.** Longitudinal relative abundances of the 40 most variable resolved microbial genus-level features across stool samples T0–T10 are shown. Taxonomic features resolving to the same genus were aggregated before analysis, and unresolved or uninformative taxonomic assignments were excluded. Genera were selected according to longitudinal variability and standardized independently across time; colors therefore represent abundance relative to the longitudinal mean of each genus rather than differences in absolute abundance between genera. Genera are ordered according to the direction of change between the pre-transition and post-transition periods.

**(B) Expanded metatranscriptomic heatmap of transcriptionally active bacteria.** The 40 bacterial taxa making the greatest longitudinal contributions to the taxonomically stratified microbial transcript pool are shown. Activity was quantified as the fraction of stratified CPM, with taxa prioritized according to peak and mean transcript contribution. Non-bacterial assignments were excluded. Activity values were log-transformed and standardized independently within each taxon for visualization. In both panels, samples are displayed chronologically using the same temporal organization as Figure 2: pre-transition (T0–T4), transition (T5–T6), and post-transition (T7–T10). Dashed vertical lines delimit the transition interval. Time is expressed as months from the first stool collection (T0 = month 0). The expanded heatmaps demonstrate that the microbial restructuring observed in Figure 2 is evident across a broader set of bacterial features and is not restricted to the 15 taxa displayed in the main figure.

**Supplementary Figure S2B. Expanded transcriptionally active bacterial community**

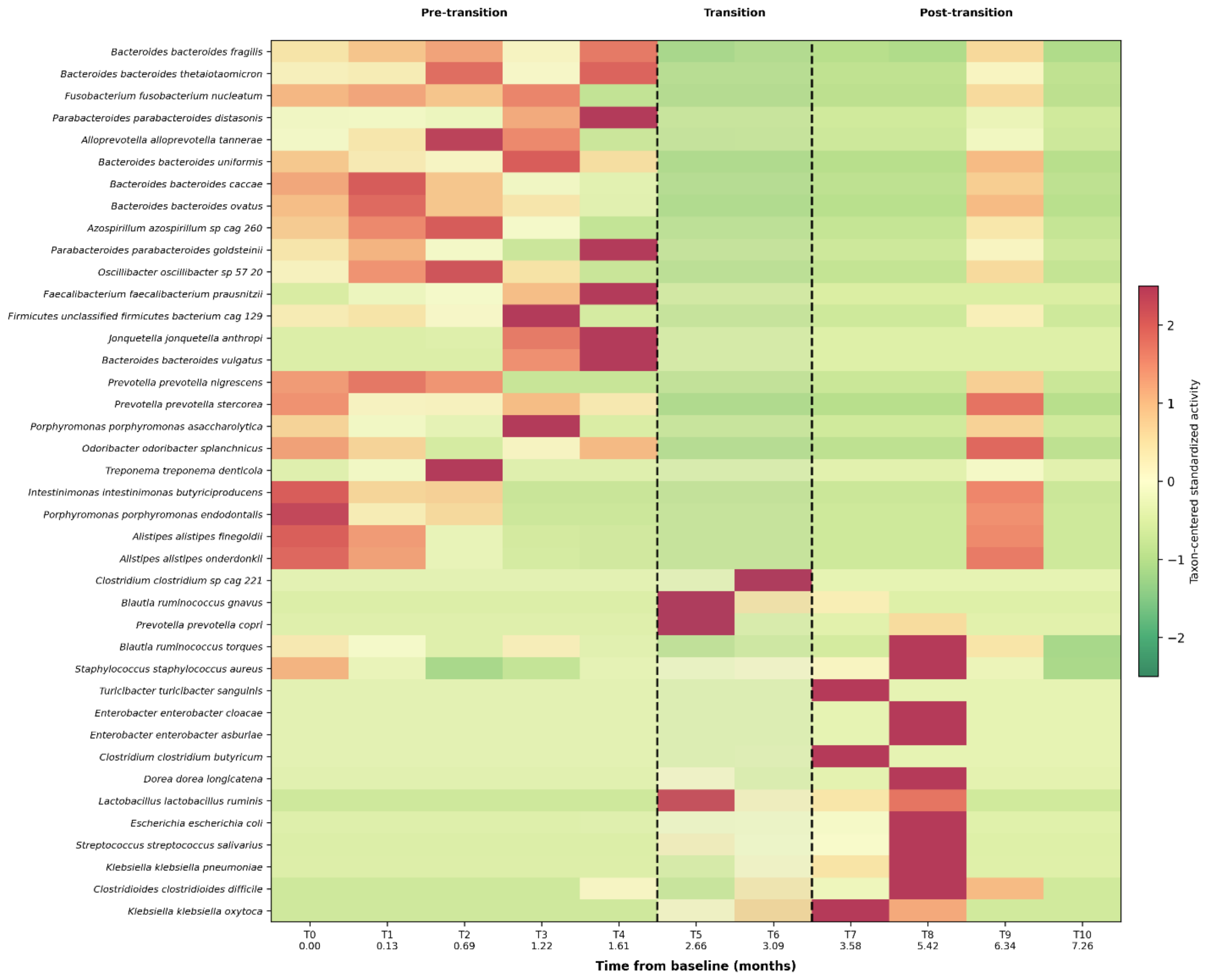

**Supplementary Figure S3. Representative longitudinal relationships between bacterial abundance and transcriptional activity.**

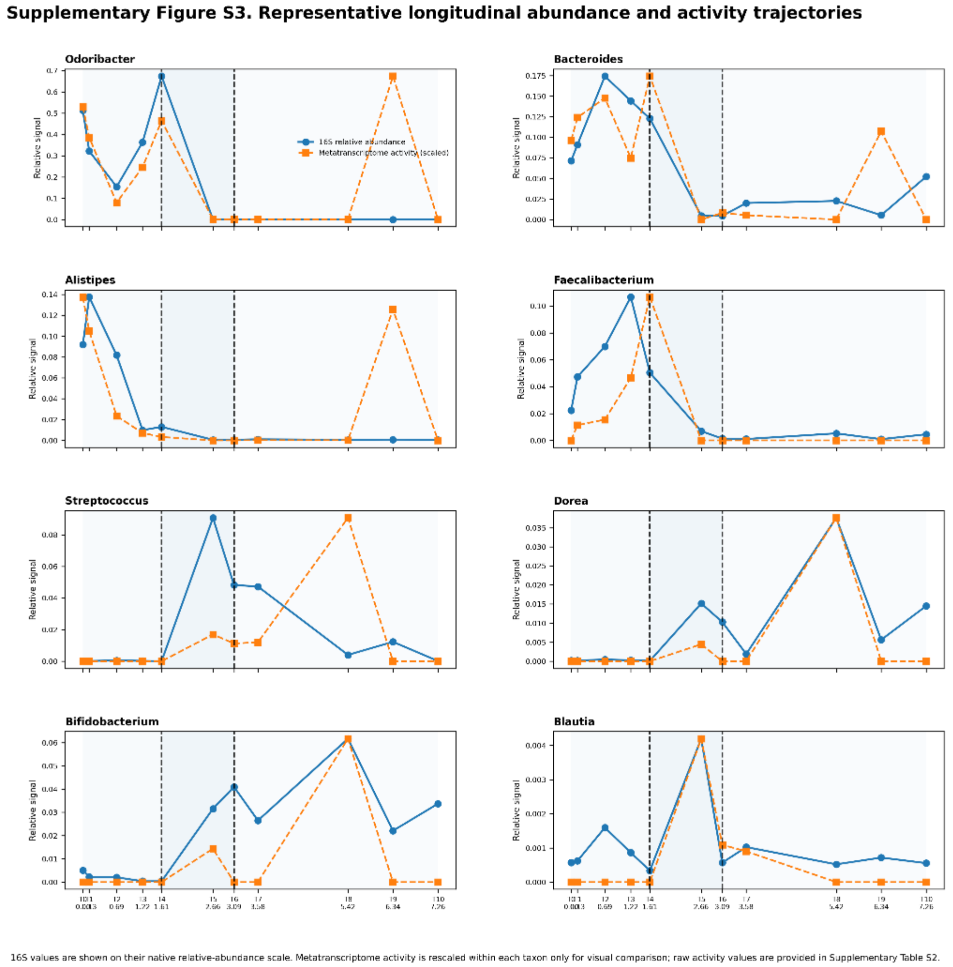


Longitudinal trajectories are shown for representative bacterial genera for which corresponding metatranscriptomic activity could be identified. Solid lines with circles indicate genus-level relative abundance measured by 16S rRNA sequencing. Dashed lines with squares indicate transcriptional activity of the dominant corresponding metatranscriptomic taxon. Representative taxa were selected on the basis of longitudinal abundance/activity and variability within the microbial datasets and not on the basis of clinical outcome. Because 16S relative abundance and metatranscriptomic transcript fractions have different numerical scales, metatranscriptomic activity was rescaled within each taxon for visualization in this figure only; Original genus-level 16S relative-abundance and taxon-attributed metatranscriptomic measurements are provided in Supplementary Tables S2 and S3, respectively. Shading and dashed vertical boundaries denote the pre-transition (T0–T4), transition (T5–T6), and post-transition (T7–T10) periods. Time is expressed as months from T0. These trajectories illustrate that taxonomic abundance and transcriptional activity provide related but non-equivalent measures of the intestinal microbial ecosystem.

**Supplementary Figure S4. Concordance of bacterial abundance and transcriptional activity and sensitivity of the microbial transition to temporal boundary definition.**

**(A) Matched longitudinal 16S abundance and metatranscriptomic activity trajectories.** Bacterial genera with identifiable metatranscriptomic counterparts and longitudinal variation in both data layers were compared across T0–T10. For visualization of trajectory shape, abundance and activity measurements were standardized independently within each feature. Solid and dashed trajectories represent 16S abundance and metatranscriptomic activity, respectively. Pearson correlation coefficients (*r*) summarize temporal concordance between the paired abundance and activity trajectories and are provided descriptively because the 11 observations represent repeated longitudinal measurements from a single participant rather than independent biological replicates. The pre-transition, transition, and post-transition periods are indicated as in Figure 2.

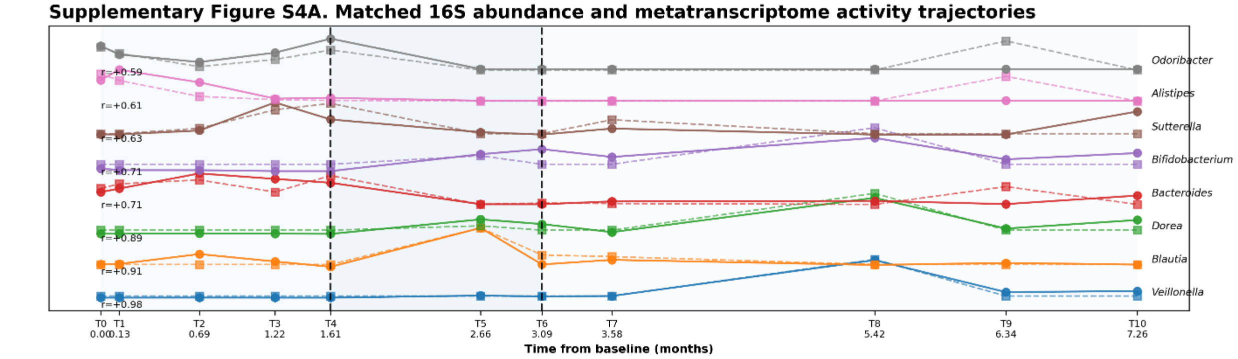


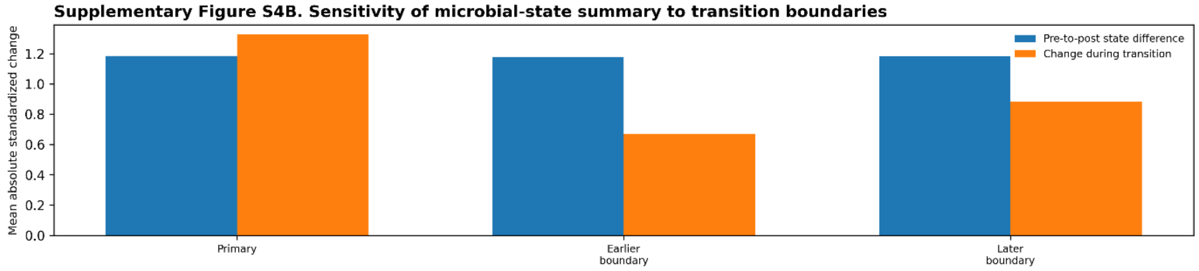


**(B) Sensitivity analysis of the microbial-state transition.** The magnitude of microbial restructuring was recalculated after shifting the temporal boundaries used to define the pre-transition, transition, and post-transition states. The primary definition (T0–T4, T5–T6, and T7–T10) was compared with earlier- and later-boundary definitions. Bars summarize the mean absolute standardized pre-to-post difference and the mean absolute change during the transition among the matched microbial features. This descriptive sensitivity analysis assesses whether the observed microbial-state restructuring remains evident under alternative temporal-boundary definitions; it does not provide independent statistical validation of the transition.

**Supplementary Figure S5. Global longitudinal remodeling of the stool metabolome.**

Heatmap showing the complete set of resolved metabolites measured longitudinally across stool samples T0–T10. In contrast to Figure 3A, which displays the 40 most longitudinally informative metabolites to facilitate biological interpretation, Supplementary Figure S5 provides a global view of the metabolomic dataset without restricting the visualization to the features selected for the main figure. Each metabolite was standardized independently across the longitudinal series; therefore, colors indicate abundance relative to the longitudinal mean of that metabolite and do not represent absolute abundance differences between metabolites. Metabolites were organized by broad biochemical categories where annotation from the metabolite name permitted classification, including purine-related, tryptophan/indole-related, sugar-acid/carbohydrate-related, amino-acid-related, and central-carbon-related metabolites; remaining metabolites were grouped as other. Rows were not hierarchically clustered, and timepoints were maintained in chronological order. Samples are divided into the same pre-transition (T0–T4), transition (T5–T6), and post-transition (T7–T10) periods used in Figures 2 and 3, with dashed vertical lines delimiting the transition interval. Time is expressed as months from T0. The expanded heatmap demonstrates that the metabolic-state transition observed in Figure 3A is evident across the broader metabolomic landscape and is not restricted to the subset of metabolites selected for the main figure. Individual metabolite identities and unstandardized measurements are provided in Supplementary Table S6.

**Supplementary Figure S5. Global longitudinal metabolomic landscape**

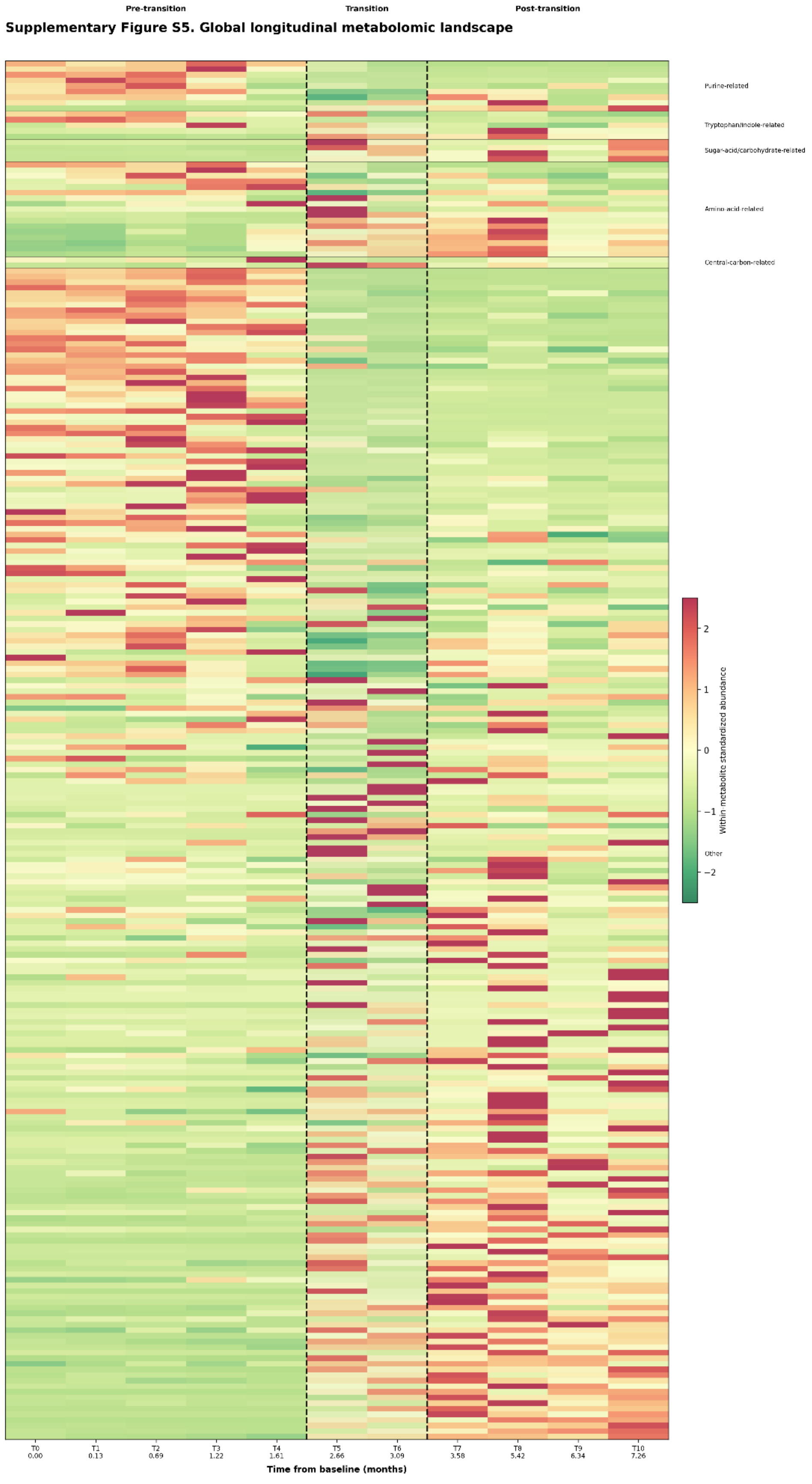

**Supplementary Figure S6. Longitudinal remodeling of individual lipid species within the major lipid classes.**

Heatmap showing individual lipid species belonging to the four lipid classes summarized in Figure 3C: ceramides, sphingomyelins (SM), lysophosphatidylcholines (LPC), and phosphatidylethanolamines (PE). Lipid species were assigned to classes from their annotated lipid names and grouped explicitly by lipid class rather than by unsupervised hierarchical clustering. Within each class, lipid species were ordered according to the direction of their longitudinal pre-to-post change. Each lipid species was standardized independently across T0–T10; consequently, colors represent changes relative to the longitudinal mean of each lipid and should not be interpreted as differences in absolute abundance between lipid species. Timepoints remain in chronological order and are divided into pre-transition (T0–T4), transition (T5–T6), and post-transition (T7–T10) periods, with dashed vertical lines identifying the transition boundaries. Time is expressed as months from T0. This species-level analysis demonstrates the extent to which the class-level trajectories shown in Figure 3C reflect coordinated remodeling among multiple constituent lipid species rather than changes driven solely by individual high-intensity lipids. The complete lipidomics matrix and lipid-class assignments are provided in Supplementary Tables S7 and S9. Original lipid measurements were provided as internal-standard-normalized relative intensities. Individual lipid species were subsequently standardized within feature across T0–T10 for heatmap visualization. Accordingly, colors represent relative longitudinal changes within each lipid species rather than absolute concentrations or directly comparable molar abundances across lipid classes.

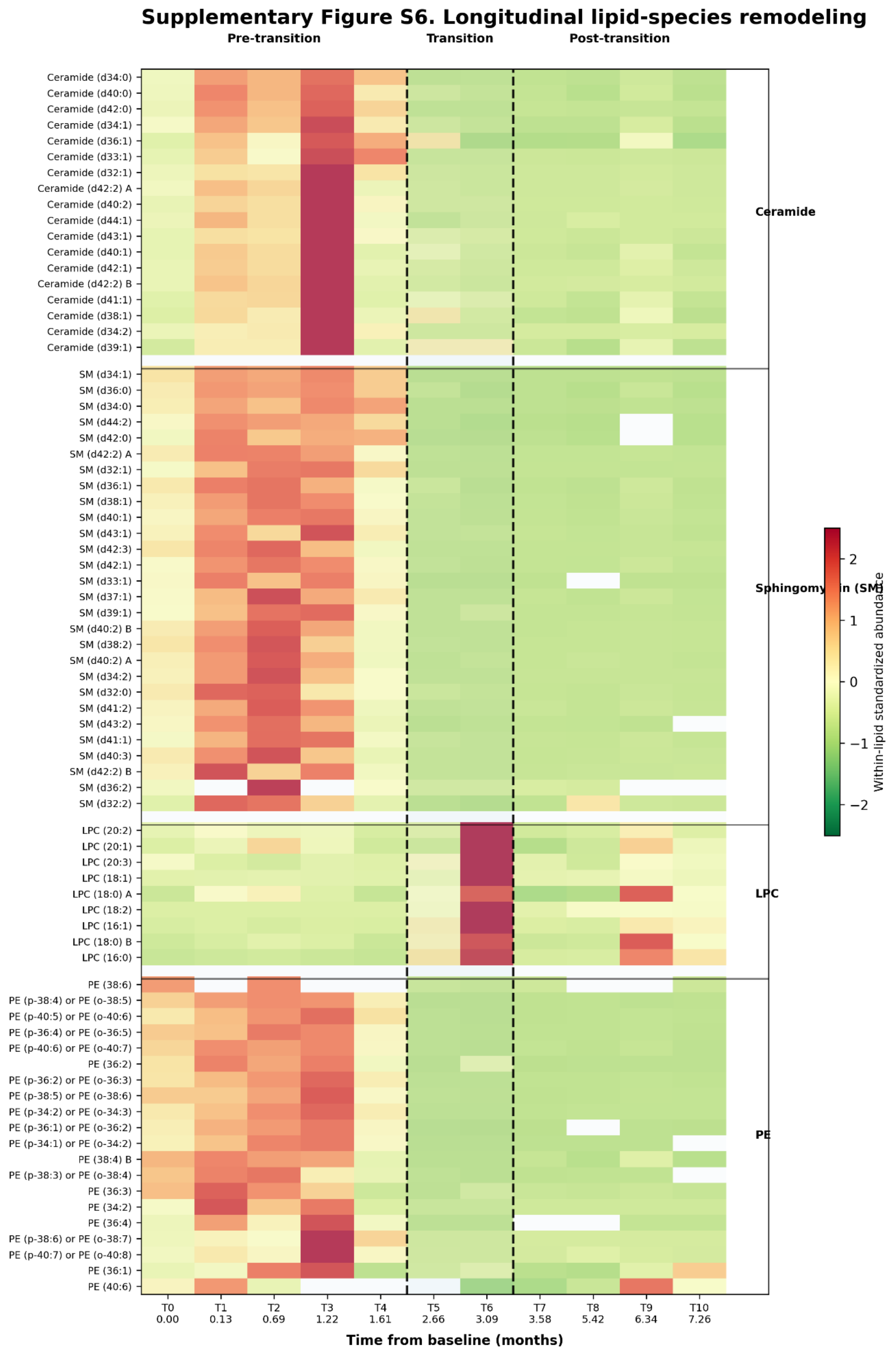
Supplementary Figure S6. Longitudinal lipid-species remodeling
Pre-transition
Transition
Post-transition
Ceramide (d34:0)
Ceramide (d40:0)
Ceramide (d42:0)
Ceramide (d34:1)
Ceramide (d36:1)
Ceramide (d33:1)
Ceramide (d32:1)
Ceramide (d42:2) A
Ceramide (d40:2)
Ceramide (d44:1)
Ceramide (d43:1)
Ceramide (d40:1)
Ceramide (d42:1)
Ceramide (d42:2) B
Ceramide (d41:1)
Ceramide (d38:1)
Ceramide (d34:2)
Ceramide (d39:1)
SM (d34:1)
SM (d36:0)
SM (d34:0)
SM (d44:2)
SM (d42:0)
SM (d42:2) A
SM (d32:1)
SM (d36:1)
SM (d38:1)
SM (d40:1)
SM (d43:1)
SM (d42:3)
SM (d42:1)
SM (d33:1)
SM (d37:1)
SM (d39:1)
SM (d40:2) B
SM (d38:2)
SM (d40:2) A
SM (d34:2)
SM (d32:0)
SM (d41:2)
SM (d43:2)
SM (d41:1)
SM (d40:3)
SM (d42:2) B
SM (d36:2)
SM (d32:2)
LPC (20:2)
LPC (20:1)
LPC (20:3)
LPC (18:1)
LPC (18:0) A
LPC (18:2)
LPC (16:1)
LPC (18:0) B
LPC (16:0)
PE (38:6)
PE (p-38:4) or PE (o-38:5)
PE (p-40:5) or PE (o-40:6)
PE (p-36:4) or PE (o-36:5)
PE (p-40:6) or PE (o-40:7)
PE (36:2)
PE (p-36:2) or PE (o-36:3)
PE (p-38:5) or PE (o-38:6)
PE (p-34:2) or PE (o-34:3)
PE (p-36:1) or PE (o-36:2)
PE (p-34:1) or PE (o-34:2)
PE (38:4) B
PE (p-38:3) or PE (o-38:4)
PE (36:3)
PE (34:2)
PE (36:4)
PE (p-38:6) or PE (o-38:7)
PE (p-40:7) or PE (o-40:8)
PE (36:1)
PE (40:6)
Ceramide
Sphingomyelin (SM)
LPC
PE
Within-lipid standardized abundance
2
1
0
−1
−2
T0 0.00
T1 0.13
T2 0.69
T3 1.22
T4 1.61
T5 2.66
T6 3.09
T7 3.58
T8 5.42
T9 6.34
T10 7.26
Time from baseline (months)

**Supplementary Figure S7. High-resolution longitudinal reconstruction of the stool host-RNA immune landscape**

**(A) Complete longitudinal immune-population matrix.** Heatmap displays the 12 RNA-derived immune population/signature scores across every available immune timepoint (T0–T9), using the same population ordering throughout. This representation preserves the full temporal structure of the immune data and demonstrates that the major reorganization apparent at T5–T6 is not an artifact of phase averaging. Increased adaptive/T- and B-cell-associated activity and reciprocal decreases in macrophage/monocyte-associated signatures become evident during the transition, followed by further remodeling in later samples. Vertical annotations identify the biologically relevant transition boundaries.

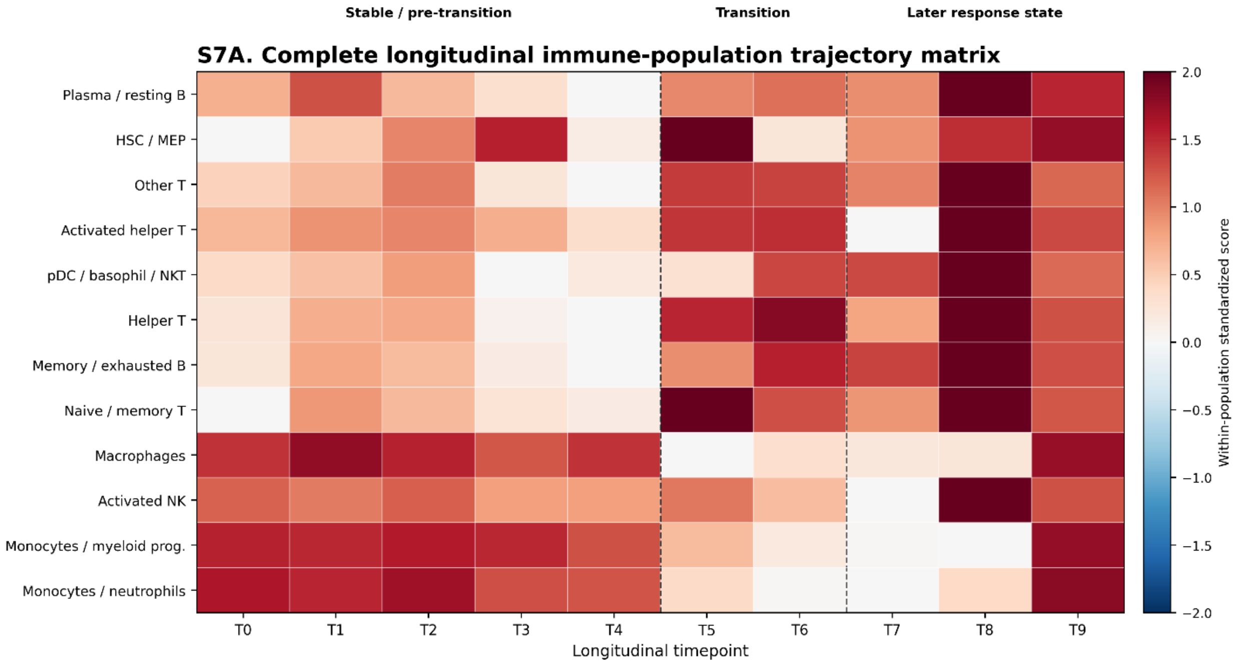


Continuous RNA-derived immune population/signature scores; T0-T9 only. Values are not direct cell counts.

**(B) Representative longitudinal immune landscapes.** Immunogram-style radial plots show four selected timepoints spanning the principal transition: T4 immediately before the transition, T5 at its onset, T6 during the subsequent transition state, and T8 during the later response-associated state. All four landscapes use identical immune-population ordering and a common radial scale, allowing direct comparison of profile geometry across time. The plots illustrate the shift from the early myeloid-prominent configuration toward increased adaptive/lymphoid-associated activity and subsequent later-state remodeling.

Immune measurements are RNA-derived signatures rather than direct enumeration of immune-cell populations. Selection of T4, T5, T6, and T8 was based on their temporal positions relative to the independently defined multi-omic transition and not on optimization of the immune profiles themselves.

**S7B. Representative longitudinal immune landscapes at the major transition timepoints**

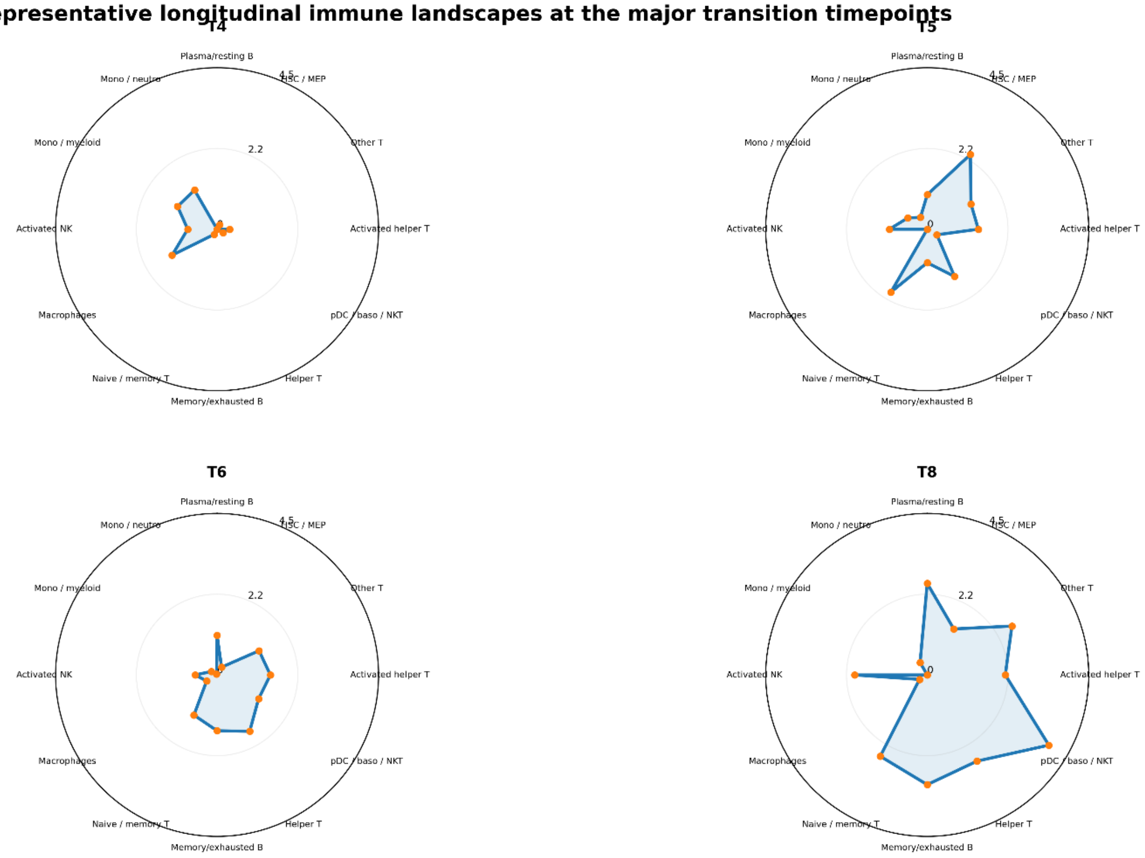


T4 = immediately before transition; T5 = Bifidobacterium-centered transition; T6 = Clostridium-centered transition; T8 = later response state. Same radial scale in all panels.

**Supplementary Figure S8. Response-aligned Immunogram atlas across eight host immune systems**. Immunogram-style radial plots summarize eight RNA-derived immune-system models reconstructed from 65 continuous longitudinal immune features: overall immune, granulocyte, monocyte, natural-killer, T-invariant, CD8 T-cell, CD4 T-cell, and B-cell systems. For visualization, features were represented using within-feature longitudinal percentile scores and aggregated into response-aligned summaries corresponding to early (T0–T4), transition (T5–T6), and later (T7–T9) intervals. The resulting profiles illustrate relative changes in immune-associated transcriptional activity across successive clinical phases rather than absolute immune-cell abundance. The original continuous measurements were retained for the quantitative multi-omic analyses in Figure 5 (Supplementary Tables S12 and S13).

The atlas reveals broad remodeling of adaptive immune programs, including T-invariant-, CD8-, CD4-, memory-, helper-, regulatory-, mucosal-, and B-cell-associated features, together with contrasting changes in monocyte/macrophage-associated programs and more heterogeneous granulocyte and natural-killer remodeling. The Immunogram atlas provides a higher-resolution response-state view complementary to the primary longitudinal analyses in Figure 4 and Supplementary Figure S7.

The response-aligned profiles are descriptive summaries of repeated longitudinal measurements from a single participant and are not comparisons among independent clinical groups. They are used to visualize concordance between the core-facility response-oriented immune framework and the independently reconstructed T0–T9 longitudinal immune trajectories. Quantitative multi-omic integration in Figure 5 used the continuous longitudinal immune measurements rather than the response-state summaries or core-facility Match p/q values.

**S8. Response-stratified reconstructed Immunogram landscapes across eight immune systems**

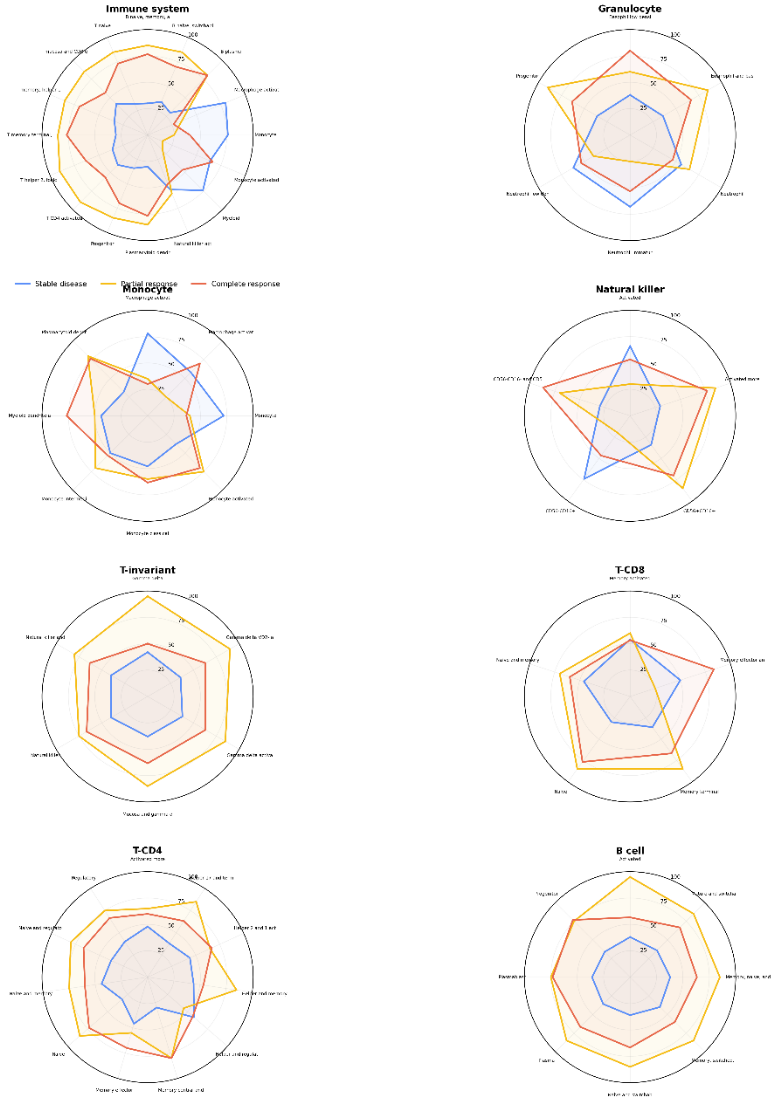


-core immune features. Each feature was converted to its longitudinal percentile (T0-T9) before phase averaging. Response-phase assignment uses the study-aligned framework: Stable T0-T4, Partial T5-T6, Complete T7-T9. The supplied core report identifies resp

**Supplementary Figure S9. Full audit trail supporting the Figure 5 multi-omic integration**

**General note.** Supplementary Figure S9 provides the molecular, chemical, and immune evidence audit supporting the integrated functional analyses in Figure 5. Functional-axis consolidation and chemical membership were established before host immune integration, using independently prioritized microbial pathways and targeted metabolomic/lipidomic associations. Historical HUMAnN pathway-to-reaction-to-UniRef90 reconstruction established patient-observed molecular support for microbial functional assignments but did not verify direct biochemical production of the associated measured chemicals. Immune associations were evaluated using continuous T0–T9 stool host-RNA-derived features. Correlations and evidence scores describe temporal concordance within a single individual and should not be interpreted as causal or population-level statistical evidence. Supporting data are provided in Supplementary Tables S14–S18.

**(A) Molecular evidence depth for the prioritized microbial functional axes.** Heatmap summarizes the evidence underlying each consolidated taxon–functional–chemical axis, including the number of contributing MetaCyc pathway units, measured chemical members, and patient-observed taxon-stratified UniRef90 gene families. Molecular support was reconstructed using the historical computational resources used by the RNA core facility (HUMAnN v3.9, UniRef90 2019_01, and MetaCyc v24). MetaCyc pathways were mapped to their constituent reactions and corresponding historical UniRef90 families, which were then intersected with taxon-stratified UniRef90 transcripts detected in the original longitudinal metatranscriptomic data. Counts represent evidence depth and should not be interpreted as independent biological replicates.

**(B) Longitudinal trajectories of the five primary microbial functional axes**. Heatmap shows standardized T0–T9 trajectories for the five nonredundant microbial states carried into the final immune integration: *Clostridium* lipid/CoA activity, *Bifidobacterium* fermentation, *Bifidobacterium* carbohydrate metabolism, *Fusobacterium* functional loss/purine remodeling, and the early-state *Bacteroides/Fusobacterium* loss/sphingomyelin-remodeling axis. Gain-side axes preserve increasing functional activity, whereas loss-side axes are oriented so that increasing axis values represent increasing loss/deactivation of the corresponding early microbial program. The trajectories illustrate the temporal separation between early-state functional loss, the T5-centered *Bifidobacterium* transition, and later *Clostridium* functional emergence.

**(C) Measured chemical members supporting each primary functional axis**. Binary membership matrix identifies the metabolite and lipid features contributing to the chemical footprint of each primary axis. Chemical members were selected during the targeted pathway–chemical analysis before immune integration and include fatty-acid and lysophospholipid species associated with the *Clostridium* lipid/CoA axis, fermentation- and carbohydrate-associated metabolites linked to the *Bifidobacterium* axes, purine-related metabolites associated with *Fusobacterium* functional loss, and sphingomyelin species marking disappearance of the early microbial state. Membership denotes inclusion in the corresponding longitudinal chemical footprint and does not imply that the measured chemical is a direct substrate or product of the associated microbial pathway.

**(D) Complete molecular-audit counts for the consolidated axes**. Table reports the pathway, chemical, pathway–chemical-pair, molecular-verification, direct-chemical-relation, semantic-mismatch, and unresolved-pair counts retained for each consolidated functional axis. These audit metrics document the successive filtering steps used to reduce the original pathway–chemical search space to the nonredundant axes used in Figure 5. A molecularly supported pathway indicates that historical pathway→reaction→UniRef90 mapping was recovered and corresponding taxon-stratified UniRef90 evidence was observed in the patient's original HUMAnN data. Absence of a verified direct chemical relationship indicates that reaction-level evidence was insufficient to classify the measured chemical as a direct substrate, product, or cofactor; such relationships were therefore retained as functional associations or state markers rather than direct biochemical mechanisms.

**(E) Feature-level immune evidence across the complete 65-feature integration**. Heatmap displays the descriptive evidence score for each of the five primary microbial functional axes against all 65 continuous RNA-derived immune features reconstructed from the eight immune-system models. The evidence score summarizes the predefined longitudinal integration criteria, including the strength of raw T0–T9 association, first-difference association, and transition-specific concordance. The two *Bifidobacterium* axes show broad evidence across multiple adaptive T-cell, mucosal/memory, regulatory, and B-cell-associated signatures, whereas the *Clostridium* and early-loss axes show more selective relationships with granulocyte-, monocyte-, myeloid-, NK-, and T-cell-associated features. Immune measurements represent stool host-RNA-derived signatures and are not direct immune-cell counts.

**General note.** Supplementary Data Figure 5 provides the audit and traceability layer underlying the integrated analyses presented in Figure 5 and Supplementary Figure S5. Microbial axes and their chemical members were defined independently of the immune data. Longitudinal associations are descriptive repeated measurements from a single individual (T0–T9); individual timepoints are not independent biological subjects, and correlation or evidence scores should not be interpreted as population-level inferential statistics. Molecular reconstruction used the historical HUMAnN/UniRef90/MetaCyc resources reported by the core facility, while direct biochemical causality was assigned only when supported by reaction-level chemical participation.

Supplementary Data Figure S9. Full audit trail for the Figure 5 multi-omic integration

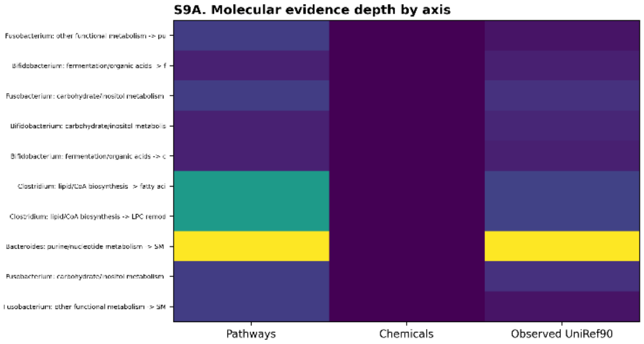


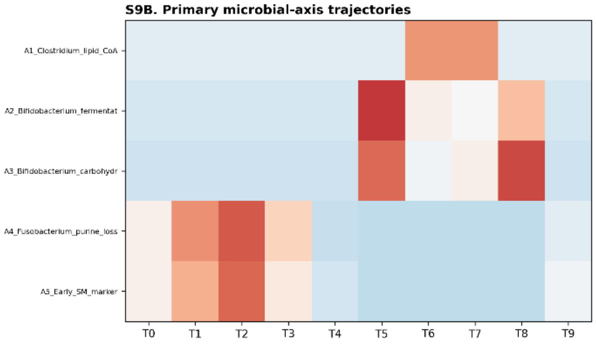


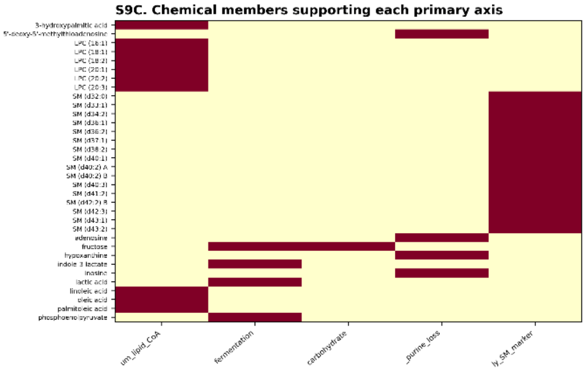


S9D. Complete molecular-audit counts

| | unique_pathways | unique_chemicals | supporting_pairs | pporting_pairs_v | t_chemical_relatio | mantic_mismatch_pa | unresolved_pairs |
|---|---|---|---|---|---|---|---|
| Fusobacterium: other functio | 2 | 0 | 5 | 3 | 0 | 0 | 5 |
| Bifidobacterium: fermentatio | 1 | 0 | 2 | 3 | 0 | 0 | 2 |
| Fusobacterium: carbohydrate/ | 2 | 0 | 3 | 5 | 0 | 0 | 3 |
| Bifidobacterium: carbohydrat | 1 | 0 | 1 | 4 | 0 | 0 | 1 |
| Bifidobacterium: fermentatio | 1 | 0 | 2 | 3 | 0 | 0 | 2 |
| Clostridium: lipid/CoA biosy | 6 | 0 | 18 | 11 | 0 | 0 | 18 |
| Clostridium: lipid/CoA biosy | 6 | 0 | 33 | 11 | 0 | 0 | 33 |
| Bacteroides: purine/nucleoti | 11 | 0 | 15 | 21 | 0 | 0 | 15 |
| Fusobacterium: carbohydrate/ | 2 | 0 | 14 | 5 | 0 | 0 | 14 |
| Fusobacterium: other functio | 2 | 0 | 20 | 3 | 0 | 0 | 20 |

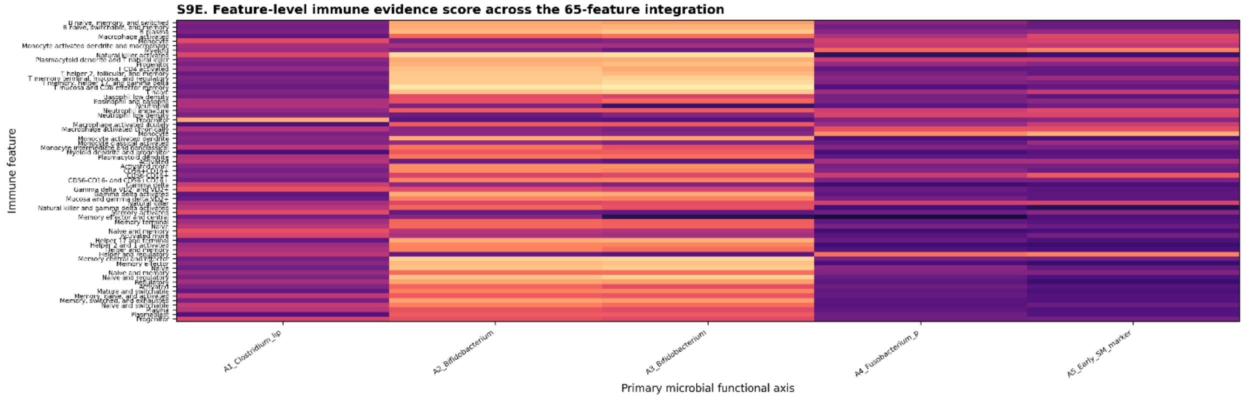
