## Supplementary Methods for "Dense Longitudinal Multi-Omics Reveals Microbial–Metabolic–Immune Succession During Immune Checkpoint Therapy"

### S1. Longitudinal clinical framework, sample alignment, and study design

#### S1.1 Longitudinal sampling and temporal coordinate system

Eleven longitudinal stool specimens (T0–T10) were collected from a single participant over approximately 7.3 months during immune checkpoint inhibitor (ICI) therapy for metastatic colon cancer (primary colon adenocarcinoma with liver metastasis). The sampling interval encompassed the early treatment period, the emergence of radiographic response, and the subsequent period of sustained tumor regression following surgical resection of the residual hepatic lesion.

To protect participant confidentiality, calendar dates were removed from publication-facing analyses. The first longitudinal stool collection was designated T0 and assigned month 0. All subsequent stool collections and clinical events were expressed as elapsed time relative to T0. Elapsed months were calculated from the corresponding interval in days using $365.25/12$ days per month.

Samples were retained according to their actual chronological sequence and temporal spacing rather than treated as equally spaced observations. The common T0-based coordinate system was used to align clinical imaging, 16S sequencing, microbial metatranscriptomics, host RNA, metabolomics, and lipidomics.

For descriptive longitudinal analyses, samples were organized into three biological periods:

- early/pre-transition: T0–T4;
- transition: T5–T6;
- later/post-transition: T7 onward.

The microbial, metabolomic, and lipidomic datasets extended through T10, whereas the host-RNA immune dataset extended through T9. No T10 immune value was interpolated or imputed. The three periods were used as descriptive temporal states and not as independent biological replicate groups. T5 and T6 were additionally retained as separate timepoints for transition-specific mechanistic analyses because substantial biological restructuring occurred within the T5–T6 interval.

#### S1.2 Longitudinal tumor-burden reconstruction

Serial computed tomography (CT) examinations were used to characterize the clinical tumor trajectory. Because complete three-dimensional volumetric segmentation was not consistently available across imaging examinations, tumor burden was estimated from the cross-sectional area of the dominant liver metastatic lesion on comparable representative coronal CT images.

Lesion boundaries were manually delineated, and enclosed image-pixel area was quantified using a consistent approach across examinations. The baseline measurement was assigned a relative tumor burden of $100\%$, and subsequent measurements were expressed relative to baseline. The resulting measurements were intended as standardized within-subject estimates of longitudinal tumor trajectory rather than formal radiologic volumetry.

The residual metastatic lesion was subsequently surgically resected. Consequently, the final $0\%$ value displayed in the tumor trajectory represents the postoperative state rather than radiographic disappearance of the lesion. Long-term follow-up imaging was displayed separately because inclusion on the treatment-period time axis would substantially compress the longitudinal observations of interest.

Clinical measurements were used to establish temporal context. Tumor response was not used to select microbial taxa, metabolites, lipids, microbial pathways, or immune features.

### S2. 16S rRNA microbiome analysis

#### S2.1 Genus-level data preprocessing

Bacterial community composition was characterized using longitudinal genus-level relative-abundance profiles derived from 16S rRNA gene sequencing. The source matrix contained multiple features sharing the same underlying genus designation but distinguished by numerical suffixes. Numerical suffixes were removed, and entries resolving to the same normalized genus were summed within each sample. Unresolved or uninformative taxonomic designations (e.g., NA, incertae_sedis, uncultured) were excluded from genus-level visualization. The resulting matrix contained one relative-abundance trajectory for each resolved bacterial genus across T0–T10.

#### S2.2 Selection of genera for the principal heatmap

Genera displayed in the principal 16S heatmap were selected using longitudinal variability calculated exclusively from the 16S dataset. For each genus, the standard deviation of relative abundance across T0–T10 was calculated, and the 15 genera with the greatest longitudinal variability were retained. No clinical-response, metabolomic, lipidomic, metatranscriptomic, or immune measurement was used in this feature-selection step.

#### S2.3 Within-genus standardization

Because genera differed substantially in absolute relative abundance, each selected genus was standardized independently across the longitudinal series:

$$\boldsymbol{z_{g,t}} = \frac{\boldsymbol{x_{g,t}} - \boldsymbol{\bar{x}_g}}{\boldsymbol{s_g}}$$

where $x_{g,t}$ is the relative abundance of genus $g$ at time $t$, $\bar{x}_g$ is its longitudinal mean, and $s_g$ is its longitudinal standard deviation. Heatmap colors therefore indicate relative longitudinal deviation within a genus and should not be interpreted as absolute abundance differences between genera. Samples were retained in chronological T0–T10 order and were not reordered by hierarchical clustering.

### S3. Microbial metatranscriptomic analysis

#### S3.1 Original sequencing-core processing

The stool metatranscriptomic data were generated using the sequencing-core computational workflow:

- KneadData: v0.7.8.1;
- Trimmomatic: v0.33;
- Host Reference Genome: *Homo sapiens* hg39 T2T Bowtie2 v0.1;
- MetaPhlAn Database: mpa_vJan25_CHOCOPhlAnSGB_202503;
- HUMAnN: v3.9;
- ChocoPhlAn: v201901_v31;
- UniRef90: 2019_01;
- MetaCyc Pathways: v24.

Taxon-stratified UniRef90 gene-family and MetaCyc pathway outputs generated by this workflow were retained for downstream analyses.

#### S3.2 Taxon-specific transcriptional activity

Taxonomically stratified microbial gene-family measurements were quantified as counts per million (CPM). Total transcriptional activity attributable to each taxon was calculated by summing all taxon-stratified UniRef90 CPM assigned to that taxon within a sample. The fraction of the total stratified microbial transcript pool attributable to taxon $i$ at time $t$ was calculated as:

$$\boldsymbol{f}_{i,t} = \frac{\mathbf{CPM}_{i,t}}{\sum_j \mathbf{CPM}_{j,t}}$$

Taxa were ranked primarily by their maximum contribution to the stratified microbial transcript pool and secondarily by their mean contribution. The 15 highest-ranking bacterial taxa were retained. Non-bacterial assignments (human, fungal, viral, bacteriophage) were excluded. Activity values used for heatmap visualization were log-transformed where required to accommodate zero/near-zero values and subsequently standardized independently within each taxon across time.

**S4. Descriptive microbial-state trajectory and temporal-boundary sensitivity analysis**

A descriptive microbial-state index was constructed to summarize the ecological transition visible independently in the 16S and metatranscriptomic datasets. Within each data layer, features were first standardized longitudinally. For each feature $i$, the difference between mean later-state and early-state standardized activity was calculated:

$$\boldsymbol{D}_i = \bar{\boldsymbol{z}}_{i,\mathbf{late}} - \bar{\boldsymbol{z}}_{i,\mathbf{early}}$$

The four features with the most negative $D_i$ values were designated early-state features, and the four with the most positive values were designated later-state features. At each timepoint, early- and later-state signatures were calculated as the unweighted mean of their constituent standardized features. A layer-specific state index was calculated as the later-state signature minus the early-state signature. The 16S and metatranscriptomic indices were then averaged to generate the descriptive trajectory.

The index was used only to summarize patterns already evident in the underlying microbial measurements and was not treated as an independent inferential endpoint. Sensitivity analyses shifted the boundaries defining early, transition, and later states (T4/T5 vs. T6/T7) to confirm that the major ecological transition did not depend on exact boundary placement.

**S5. Genus-harmonized 16S abundance–metatranscriptomic activity integration**

**S5.1 Taxonomic harmonization**

For cross-layer analyses, 16S and metatranscriptomic taxonomic labels were harmonized at the genus level. Species-level RNA assignments were collapsed to their corresponding genus, and entries sharing the same normalized genus were summed. Only genera that could be resolved consistently between the two microbial data layers were used for abundance–activity comparison.

**S5.2 Transition measures**

For each genus, the following quantities were calculated separately for 16S relative abundance and RNA activity:

- early mean: T0–T4;
- transition mean: T5–T6;
- later mean: T7–T9;
- stepwise changes: T4 → T5, T5 → T6, and T4 → T6;
- period changes: early → transition, and early → later.

Pseudocount-aware log-ratio changes were used where required for features containing zero or near-zero values.

**S5.3 Directional taxon prioritization**

Taxa were ranked separately for three biological behaviors:

1. activation/emergence: representing increasing transcriptional activity;
2. loss/deactivation: representing declining transcriptional activity;
3. overall cross-layer restructuring: a direction-neutral measure of discordance.

An absolute RNA-contribution criterion was included so that a taxon exhibiting a large fold change from a negligible baseline could not outrank an organism making a substantial contribution to the active microbial transcript pool. Taxon prioritization was completed without metabolomic, lipidomic, immune, or clinical-response information.

**S6. Taxon-resolved MetaCyc pathway analysis**

Taxon-stratified MetaCyc pathway profiles were interrogated for the prioritized gain- and loss-associated taxa. For each taxon–pathway combination, three complementary measurements were evaluated:

1. absolute taxon-attributed pathway signal;
2. fraction of total pathway signal attributable to the taxon;
3. pathway activity relative to total taxon RNA activity.

The third measurement was used to distinguish pathway-specific functional allocation from organism-wide transcriptional expansion. Pathways emerging from an undetectable baseline were classified as emergent rather than assigned an arbitrarily large fold change.

Candidate specialization was considered only when pathway and total taxon RNA measurements were sufficiently measurable for normalization.

Gain and loss were treated asymmetrically. Gain-associated taxa were evaluated according to transition-state pathway activity and contribution, whereas declining taxa were evaluated according to pre-transition functional activity and subsequent loss. Pathway evidence was classified descriptively into categories including emergent pathway activity, candidate specialization, substantial participation, loss-associated function, and non-priority/zero-baseline signal.

### S7. Metabolomics analytical platforms, metabolite annotation, normalization, and longitudinal analysis.

#### Analytical facility and platforms

Stool metabolomic and lipidomic analyses were performed at the West Coast Metabolomics Center, University of California, Davis (Fiehn Laboratory). The core facility supplied data dictionaries describing its GC–TOF MS primary-metabolism platform and HILIC–ESI–QTOF MS/MS platform for biogenic amines and other small-molecule metabolites.

#### Primary metabolism by GC–TOF MS

The core facility's primary-metabolism platform uses gas chromatography coupled to electron-ionization time-of-flight mass spectrometry. The documented method employs a Restek Rtx-5Sil MS capillary column (30 m × 0.25 mm internal diameter, 0.25 μm film thickness) with helium carrier gas. The column temperature increases from 50°C to 330°C. The documented mass spectrometer is a LECO Pegasus IV operated with electron ionization at 70 eV.

Raw chromatographic data are processed using ChromaTOF for baseline correction, peak detection, and mass spectral deconvolution. Compound identification is performed using the Fiehn Laboratory BinBase system, which combines retention-index matching, characteristic mass fragments, spectral similarity, and quality-control filters. Fatty-acid methyl ester internal standards are used for retention-index alignment.

Metabolite abundance is reported as the peak height of a selected quantification ion. The core's documented normalization framework uses the summed peak heights of identified metabolites (mTIC) to account for analytical variation. Additional quality-control samples and batch correction procedures may be employed to evaluate instrumental drift. Reported values represent relative semiquantitative abundances rather than absolute metabolite concentrations.

**Polar metabolites and biogenic amines by HILIC–QTOF MS/MS**

The core facility's HILIC platform uses ultrahigh-performance liquid chromatography coupled to electrospray-ionization quadrupole time-of-flight tandem mass spectrometry, with positive- and negative-ion acquisition as appropriate.

The documented chromatographic method uses a Waters Acquity Premier BEH Amide column (1.7 μm, 2.1 × 50 mm) at 45°C, with water/acetonitrile mobile phases containing ammonium formate and formic acid. The method employs a short gradient and a flow rate of 0.8 mL/min.

The facility's standard processing workflow uses MS-DIAL for peak detection, chromatographic alignment, and compound annotation. Metabolites are identified using an in-house mass-to-charge/retention-time library and tandem mass spectral matching against NIST and MassBank of North America (MoNA) reference libraries. MS/MS-based annotations undergo manual review.

The core reports metabolite annotation confidence according to a modified Metabolomics Standards Initiative (MSI) classification, ranging from high-confidence assignments supported by accurate mass, retention time, and MS/MS to unidentified features.

Relative metabolite abundances are reported as quantification-ion peak heights. The documented normalization approach generally uses the sum of internal-standard peak heights (iTIC), with mTIC-based normalization available under certain sample-processing conditions.

**Longitudinal metabolomic analysis**

The annotated metabolomics matrix contained 246 features measured across T0–T10. Feature names were harmonized, and duplicate normalized labels were consolidated when applicable. The original quantitative measurements were retained for individual metabolite trajectories and downstream integration.

Longitudinal variability was evaluated within the metabolomics dataset independently of microbial, immune, or clinical-response information. The 40 most variable metabolites were selected for the principal visualization. Each selected feature was standardized longitudinally to emphasize relative temporal changes rather than differences in absolute signal intensity.

Representative metabolite trajectories were displayed using the original core-provided quantitative values.

**Integration with microbial functional pathways**

Targeted chemical integration was restricted to microbial pathways independently prioritized from taxon-resolved metatranscriptomic analyses. Candidate pathway–chemical relationships were evaluated using longitudinal Pearson and Spearman correlations, first-difference correlations, and directional concordance across T4→T5, T5→T6, and T4→T6.

Chemical relationships were interpreted according to available biochemical evidence. Direct substrate, product, or cofactor assignments required reaction-level support, whereas chemically indirect relationships were retained as functional associations or transition-state markers.

Because the mass-spectrometric measurements were relative semiquantitative signals, pathway–chemical associations were not interpreted as direct measurements of metabolite production rates, substrate consumption, or metabolic flux.

#### S7.1 Source matrix and preprocessing

The quantitative known-metabolite dataset contained 246 annotated features across T0–T10. Measurements were converted to numeric values. Features lacking sufficient longitudinal observations or variation were excluded from trajectory calculations. When duplicate normalized metabolite labels were present, measurements were averaged to produce a single longitudinal feature. Original quantitative measurements were retained as the authoritative metabolomics matrix.

#### S7.2 Scale-aware longitudinal variability

To select an interpretable subset for visualization without allowing absolute measurement scale to dominate selection, longitudinal variability was estimated using the median absolute deviation relative to the median absolute signal. Standard deviation was used as a fallback for features with median values approaching zero. The 40 metabolites with the highest variability scores were retained for global heatmaps. Feature selection used metabolomics measurements alone. Each selected metabolite was standardized longitudinally:

$$z_{i,t} = \frac{x_{i,t} - \bar{x}_i}{s_i}$$

#### S7.3 Representative metabolites

Representative trajectories were displayed for inosine, uric acid, tryptophan, indole-3-lactate, galactonic acid, and xylonic acid isomer. These trajectories used original core-provided semiquantitative measurement scales. These metabolites were chosen to represent prominent biochemical domains evident in the metabolomics data and were not selected according to tumor-response correlation.

**S8. Stool lipidomics analysis**

**S8.1. Stool lipid extraction and LC-HRMS/MS acquisition**

Stool lipidomic profiling was performed at the University of California, Davis West Coast Metabolomics Center according to study-specific protocol MX 378087. Approximately 4 mg of each stool specimen was subjected to lipid extraction using the Matyash liquid–liquid extraction procedure with methyl tert-butyl ether (MTBE), methanol, and water {Matyash, 2008 #30}. The upper organic phase containing extracted lipids was collected and dried before analysis.

Dried lipid extracts were reconstituted in 110 µL of methanol/toluene (9:1, v/v) containing 50 ng/mL CUDA. Samples were shaken for 20 seconds, sonicated for 5 minutes at room temperature, and centrifuged at 16,100 × g for 2 minutes. Extracts were divided into aliquots, including 33 µL transferred to glass-insert vials for positive- and negative-ion lipidomics, with a remaining aliquot reserved for pooled sample preparation.

Chromatographic separation was performed using an Agilent 1290 Infinity LC system equipped with a Waters ACQUITY UPLC CSH C18 column (1.7 µm, 2.1 × 100 mm) and a VanGuard precolumn (2.1 × 5 mm). The mobile-phase gradient was programmed as follows: 15% solvent B at 0 min, 30% B at 2 min, 48% B at 2.5 min, 82% B at 11 min, 99% B at 11.5–12 min, and return to 15% B at 12.1 min, followed by equilibration until 15 min. The flow rate was 0.6 mL/min.

Positive-ion electrospray analysis was performed using an Agilent 6550 high-resolution mass spectrometer, with a 5 µL injection volume, an acquisition range of m/z 120–1200, and an acquisition rate of 2 spectra/s. Negative-ion electrospray analysis was performed using an Agilent 6530 QTOF mass spectrometer under the same chromatographic conditions, with an acquisition range of m/z 60–1200 and an acquisition rate of 2 spectra/s. The core-facility documentation reported resolving powers of approximately 10,000 and 30,000 for positive- and negative-ion acquisition, respectively.

**S8.2. Lipidomic data preprocessing, annotation, and class-level analysis**

The UC Davis West Coast Metabolomics Center supplied annotated lipid-species measurements as internal-standard-normalized relative intensities. These measurements were used for longitudinal analyses without conversion to absolute molar concentrations.

Lipid identifiers were harmonized, and duplicate normalized labels were averaged when present. Measurements were converted to numeric values, and features with insufficient longitudinal observations or no measurable variation were excluded from trajectory calculations.

Lipid species were assigned to four principal classes using explicit annotation-based rules: ceramides (Cer), sphingomyelins (SM), lysophosphatidylcholines (LPC), and phosphatidylethanolamines (PE). The complete lipidomic measurement matrix and class assignments are provided in Supplementary Tables S7 and S9, respectively.

For class-level visualization, each lipid species was independently standardized across the longitudinal series (T0–T10):

$$z_{i,t} = \frac{x_{i,t} - \bar{x}_i}{s_i}$$

where ($x_{i,t}$) represents the internal-standard-normalized relative intensity of lipid species ($i$) at timepoint ($t$), ($\overline{x}_i$) is its longitudinal mean, and ($s_i$) is its longitudinal standard deviation.

The standardized class-level trajectory was calculated as the arithmetic mean of the standardized constituent species:

$$L_{c,t} = \frac{1}{n_c} \sum_{i \in c} z_{i,t}$$

where ($L_{c,t}$) represents the standardized trajectory of lipid class ($c$) at timepoint ($t$), and ($n_c$) is the number of lipid species assigned to that class.

This procedure summarizes coordinated longitudinal behavior among lipid species while preventing high-intensity species from disproportionately determining the class trajectory. The resulting class scores are dimensionless and should not be interpreted as absolute lipid-class concentrations.

**S8.3. Lipid-class signal ratios**

Ceramide/sphingomyelin (Cer/SM) and lysophosphatidylcholine/phosphatidylethanolamine (LPC/PE) ratios were calculated at each timepoint using summed original internal-standard-normalized relative intensities, before longitudinal standardization:

$$R_{\mathrm{Cer/SM},t} = \frac{\sum_{i\in\mathrm{Cer}} x_{i,t}}{\sum_{j\in\mathrm{SM}} x_{j,t}}$$

$$R_{\mathrm{LPC/PE},t} = \frac{\sum_{i\in\mathrm{LPC}} x_{i,t}}{\sum_{j\in\mathrm{PE}} x_{j,t}}$$

Ratios of standardized values were not used. Because the original measurements represent relative analytical signals rather than calibrated molar concentrations, these ratios describe relative changes in lipid-class signal relationships and do not establish absolute molar ratios, enzymatic conversion rates, or metabolic flux.

The lipid-class trajectories and ratio calculations support Figure 3C,D and Supplementary Figure S6. The underlying quantitative measurements, lipid-class assignments, and ratio calculations are documented in Supplementary Tables S7, S9, and S10.

### S9. Host-RNA immune analysis and quality control

#### S9.1 Twelve-population immune representation

Host-derived RNA recovered from stool was analyzed separately from microbial RNA. The initial lower-dimensional immune representation contained 12 RNA-derived population/signature scores: plasma/resting B, HSC/MEP, other T, activated helper T, pDC/basophil/NKT, helper T, memory/exhausted B, naïve/memory T, macrophage, activated NK, monocyte/myeloid progenitor, and monocyte/neutrophil. Immune measurements were available across T0–T9. No T10 immune measurement was imputed or interpolated. These measurements represent transcriptionally inferred population state signatures rather than direct cell counts.

#### S9.2 Phase-level immune analysis

Phase means were calculated for Early (T0–T4), Transition (T5–T6), and Later (T7–T9). The immediate transition-associated change for each population was calculated as:

$$\boldsymbol{\Delta_i = \overline{x}_{i,\mathrm{T5:T6}} - x_{i,\mathrm{T4}}}$$

#### S9.3 Descriptive immune composites

The adaptive/T-associated composite was calculated as the unweighted mean of *activated helper T*, *helper T*, *naïve/memory T*, and *other T*. The myeloid-associated composite was calculated as the unweighted mean of *macrophage*, *monocyte/myeloid progenitor*, and *monocyte/neutrophil*. These composites served purely as descriptive summaries.

#### S9.4 Comprehensive core-facility immune audit

Before Figure 5 integration, the complete RNA-core immune archive was independently reviewed. Continuous T0–T9 values were reconstructed for 65 immune features across eight immune models:

1. overall immune system;
2. granulocyte system;
3. monocyte system;
4. natural-killer system;
5. T-invariant system;
6. T-CD8 system;
7. T-CD4 system;
8. B-cell system.

Percentile/rank-transformed Bubble/Immunogram values were used strictly for quality control and provenance visualization, whereas quantitative multi-omic integrations used continuous unadjusted feature values.

#### S10. Historical HUMAnN pathway–reaction–UniRef90 reconstruction

Molecular verification was performed using the exact historical database resources reported by the sequencing core:

- HUMAnN: v3.9;
- UniRef90: 2019_01;
- MetaCyc: v24;
- Reaction mapping: metacyc_reactions_level4ec_only.uniref;
- Structured pathways: metacyc_pathways_structured_filtered_v24_subreactions.

For each prioritized MetaCyc pathway, the structured pathway definition was used to decompose the pathway into constituent reactions. Historical reaction assignments were mapped to UniRef90 families using the corresponding UniRef90-to-MetaCyc reaction mapping resource. These families were intersected with taxon-stratified UniRef90 families detected in the original patient metatranscriptomic outputs.

A pathway was considered molecularly supported when:

1. the historical pathway definition contained relevant reactions;
2. the reaction mapped to historical UniRef90 families; and
3. corresponding taxon-stratified UniRef90 families were actively transcribed in the patient's data.

This established the chain:

$$\text{MetaCyc pathway} \longrightarrow \text{reaction} \longrightarrow \text{UniRef90} \longrightarrow \text{patient taxon RNA}$$

Molecular pathway support was explicitly distinguished from direct pathway–chemical evidence. A measured chemical was classified as a direct biochemical participant only when the reaction-level mapping supported its role as substrate, product, or cofactor; otherwise, it was classified as a state marker.

### S11. Targeted microbial pathway–chemical–immune integration

#### S11.1 Prespecified biological hypotheses

Chemical integration was restricted to pathways supported by the preceding taxon-resolved functional analysis across five biological domains:

1. *Clostridium* → inositol/carbohydrate metabolism;
2. *Clostridium* → purine metabolism;
3. *Clostridium* → fatty-acid/CoA metabolism;
4. *Bifidobacterium* → carbohydrate fermentation;
5. Loss of baseline *Bacteroides/Fusobacterium/Odoribacter* functions.

#### S11.2 Longitudinal association measures and transition SD shifts

For each candidate pathway–chemical pair, three complementary association metrics were calculated across the longitudinal series:

1. Continuous Trajectory Correlations: Parametric Pearson correlation ($r$) and non-parametric Spearman rank correlation ($\rho$) were evaluated on unstandardized and $z$-standardized continuous measurements across T0–T10.
2. First-Difference Trajectory Correlations: First differences ($\Delta x_t = x_t - x_{t-1}$) were computed for both pathway RNA signal and chemical intensity across consecutive timepoints. Pearson and Spearman correlations were then evaluated on the resulting difference vectors (ΔPathway vs. ΔChemical) to control for autocorrelation.

3. Transition-Specific Directional Concordance and SD Shifts: Stepwise directional shifts were quantified for three transition intervals (T4 → T5, T5 → T6, and T4 → T6). To quantify transition magnitudes across different measurement scales, standard deviation shifts ($\Delta z_{\text{transition}}$) were computed as:

$$\boldsymbol{\Delta z}_{\textbf{transition}} = \frac{\boldsymbol{x}_{t_2} - \boldsymbol{x}_{t_1}}{\boldsymbol{s}_{\textbf{longitudinal}}}$$

where $x_{t_1}$ and $x_{t_2}$ represent raw values at the boundary timepoints and $s_{\text{longitudinal}}$ represents the standard deviation of the feature across the entire series.

S11.3 Consolidation into nonredundant functional axes and host immune integration

Redundant pathway–chemical relationships were consolidated into five nonredundant functional axes:

1. *Clostridium* lipid/CoA activity with fatty-acid/LPC remodeling;
2. *Bifidobacterium* fermentation with fermentation-associated chemicals;
3. *Bifidobacterium* carbohydrate metabolism with carbohydrate-associated metabolites;
4. *Fusobacterium* functional loss with purine remodeling;
5. Loss of the early *Bacteroides/Fusobacterium* state with sphingomyelin remodeling.

These functional axes were integrated with the 65 continuous host-derived stool immune features across T0–T9. Association strength was quantified using raw longitudinal Pearson/Spearman correlations, first-difference correlations, and directional shifts across T4 → T5 and T5 → T6. Loss-associated functional axes were inverted such that higher values reflected increasing functional loss/deactivation. Final temporal-mechanistic consolidation compared microbial function, chemical footprints, and host immune signatures separately across the T4 → T5 and T5 → T6 transition windows.
