## Supplementary Data Tables 1-18 for "Dense Longitudinal Multi-Omics Reveals Microbial–Metabolic–Immune Succession During Immune Checkpoint Therapy": Supplementary_Data_Tables_Legends.docx

**Supplementary Data and Table Legends**

**Supplementary Table S1. Immune checkpoint inhibitor treatment administration schedule.**

De-identified schedule of nivolumab (anti-PD-1) and ipilimumab (anti-CTLA-4) administrations during treatment of colorectal cancer metastatic to the liver. Treatment times are expressed as elapsed days and months relative to the first stool collection (T0 = day 0), which preceded initiation of immunotherapy by 17 days. Nivolumab was administered at approximately 3 mg/kg every two weeks for 16 documented administrations, with concurrent ipilimumab at approximately 1 mg/kg every six weeks for six administrations. Calendar dates and patient-specific absolute doses are omitted to reduce re-identification risk. The schedule supports the treatment timeline in Figure 1A.

**Supplementary Table S2. Complete longitudinal genus-level 16S rRNA relative-abundance matrix.**

Genus-level bacterial relative-abundance measurements across 11 longitudinal stool samples (T0–T10). Taxonomic assignments resolving to the same genus were aggregated after harmonization of numerical suffixes, and unresolved taxonomic designations were excluded. The table retains the complete resolved genus-level matrix used for the microbial community analyses in Figure 2A, including genera not selected for the principal heatmap. Values represent relative bacterial abundance rather than absolute microbial biomass.

**Supplementary Table S3. Complete longitudinal taxon-stratified microbial transcriptional activity matrix.**

Taxon-attributed microbial RNA activity across T0–T10, derived from taxon-stratified UniRef90 gene-family measurements generated by the HUMAnN metatranscriptomic workflow. Measurements are expressed as counts per million (CPM) or corresponding taxon contributions to the stratified microbial transcript pool, as specified in the workbook. The complete matrix supports the transcriptional activity heatmap in Figure 2B and subsequent abundance–activity comparisons. RNA activity represents taxon-attributed transcriptional signal rather than transcription per bacterial cell.

**Supplementary Table S4. Microbial feature selection, state-index construction, and temporal-boundary sensitivity analysis.**

Supporting calculations for the descriptive longitudinal microbial-state analysis in Figure 2C. The workbook documents selection of longitudinally variable bacterial taxa, identification of early- and later-state features, and construction of standardized microbial-state signatures from 16S and metatranscriptomic measurements. Additional worksheets provide the alternative temporal-boundary schemes used to assess the stability of the observed ecological transition. The index is descriptive and was not optimized against clinical response, metabolomic, lipidomic, or immune measurements.

**Supplementary Table S5. Genus-harmonized directional ranking of bacterial abundance and transcriptional activity.**

Longitudinal 16S relative-abundance profiles and taxon-stratified UniRef90 metatranscriptomic activity were harmonized at genus level and evaluated independently of clinical response and other omics measurements. Separate worksheets provide rankings for transcriptional activation/emergence (S5A), loss/deactivation (S5B), and overall direction-neutral cross-layer restructuring (S5C). Ranking components include longitudinal abundance and RNA changes, transition-specific activity, abundance–activity discordance, and absolute RNA contribution to prevent disproportionate prioritization of taxa with negligible transcriptional activity. S5D provides harmonization and ranking-run audit metrics. Original ranking scores and eligibility classifications are preserved without recalculation. These analyses support Figure 2D and the initial taxon prioritization for Figure 5.

**Supplementary Table S6. Complete longitudinal stool metabolomics matrix.**

Quantitative measurements for 246 annotated stool metabolites across T0–T10, as provided by the UC Davis West Coast Metabolomics Center and harmonized for longitudinal analysis. The table includes all retained metabolite features, irrespective of their selection for the principal heatmap or individual trajectory plots. Original core-provided quantitative measurements are preserved to support Figure 3A,B and subsequent targeted pathway–chemical integration. Values represent relative semiquantitative metabolomic measurements rather than absolute molar concentrations.

**Supplementary Table S7. Complete longitudinal stool lipidomics matrix.**

Individual lipid-species measurements across T0–T10 generated by the UC Davis West Coast Metabolomics Center using reverse-phase LC-HRMS analysis. Values represent internal-standard-normalized relative lipid intensities. The complete matrix includes the measured lipid species used for longitudinal analyses, including ceramides, sphingomyelins, lysophosphatidylcholines, phosphatidylethanolamines, and other annotated lipid classes. Measurements were retained in their core-provided normalized form for subsequent class-level and functional integration analyses.

**Supplementary Table S8. Longitudinal metabolite variability ranking and feature-selection audit.**

Supporting data for selection of the 40 most longitudinally variable metabolites displayed in Figure 3A. Metabolites were ranked using a scale-aware longitudinal variability measure calculated independently of clinical response and other omics measurements. The table provides feature identifiers, variability scores, selection status, and heatmap ordering. Standardized trajectories used for visualization are distinguished from the original quantitative metabolite measurements provided in Supplementary Table S6.

**Supplementary Table S9. Lipid-species classification and longitudinal lipid-class trajectory audit.**

Assignment of individual lipid species to the principal lipid classes used in Figure 3C, including ceramides, sphingomyelins (SM), lysophosphatidylcholines (LPC), and phosphatidylethanolamines (PE). The table documents lipid identifiers, class membership, and the constituent species used to construct class-level trajectories. Each species was standardized independently across the longitudinal series before averaging within its assigned class. These class trajectories summarize coordinated changes in relative lipid signals and are not direct measurements of lipid synthesis or conversion rates.

**Supplementary Table S10. Longitudinal ceramide/sphingomyelin and lysophosphatidylcholine/phosphatidylethanolamine signal ratios.**

Supporting calculations for the Cer/SM and LPC/PE trajectories shown in Figure 3D. For each timepoint, the internal-standard-normalized relative intensities of lipid species belonging to each class were summed before calculation of the corresponding class ratio. Ratios were calculated from unstandardized core-provided measurements, not from within-species z-scores. Because the lipidomics data represent relative analytical intensities rather than calibrated molar concentrations, these ratios describe relative class-level signal relationships and should not be interpreted as absolute molar ratios or direct biochemical conversion rates.

**Supplementary Table S11. Complete longitudinal 12-population stool host-RNA immune-signature matrix.**

RNA-derived scores for 12 immune population/state signatures across T0–T9, supporting Figure 4A–C and Supplementary Figure S7. The matrix preserves the longitudinal measurements used to characterize adaptive/lymphoid- and myeloid-associated immune remodeling. The immune scores represent transcriptionally inferred population or state signatures rather than direct cell counts. No T10 immune measurement was available or imputed.

**Supplementary Table S12. High-resolution longitudinal host immune-feature measurements and metadata.** Continuous stool host-RNA-derived scores for 65 immune features across T0–T9, organized into eight immune-system models: overall immune, granulocyte, monocyte, natural-killer, T-invariant, CD8 T-cell, CD4 T-cell, and B-cell systems. S12A contains the original continuous measurements, S12B the within-feature standardized trajectories, S12C feature identifiers and model membership, and S12D descriptive phase and transition summaries. These measurements supported the eight-model immune analyses in Figure 4 and the quantitative microbial–chemical–immune integration in Figure 5. Scores represent transcriptionally inferred immune-associated states rather than direct cell counts. No T10 immune measurement was available or imputed.

**Supplementary Table S13. Eight-model immune-system summaries and response-aligned profiles.**

Longitudinal and response-aligned summaries of the eight RNA-core immune-system models, supporting Figure 4D and Supplementary Figure S8. Model-level summaries were derived from constituent continuous immune features and organized according to the early, transition, and later response-aligned intervals. The table retains the values used to reconstruct the eight-system Immunogram representation. These summaries describe repeated observations from a single individual and are not comparisons among independent clinical-response groups.

**Supplementary Table S14. Consolidation and pathway membership of microbial functional axes before immune integration.**

Taxon-associated MetaCyc pathways were evaluated against measured stool metabolites and lipids using longitudinal correlations, first-difference correlations, and biologically expected transition-direction concordance. Of 9,400 evaluated pathway–chemical pairs, 327 met the predefined evidence criteria and were organized into 44 consolidated functional axes. Ten core axes were retained before host immune integration, supported by 113 pathway–chemical pairs comprising 34 axis-specific pathway assignments and 23 distinct MetaCyc pathways. The workbook provides core-axis rankings, pathway–chemical membership, complete consolidated-axis definitions, evidence classifications, and selection methodology. Associated-state-marker axes are distinguished from stronger functional relationships; these classifications do not establish direct biochemical production of measured chemicals. Axis consolidation was performed independently of host immune measurements and clinical outcome.

**Supplementary Table S15. Historical MetaCyc pathway–reaction–UniRef90 molecular-verification audit.**

Molecular evidence for ten prioritized microbial functional axes was reconstructed using the historical HUMAnN v3.9, UniRef90 2019_01, and MetaCyc v24 resources employed in the original metatranscriptomic analysis. The workbook contains axis-level molecular-support classifications (S15A), pathway–reaction verification (S15B), detailed reaction-associated UniRef90 evidence (S15C), taxon-stratified UniRef90 family measurements across T4–T6 (S15D), original family-level observations (S15E), run-summary statistics (S15F), and verification methodology (S15G). All ten axes exhibited patient-observed molecular support and were classified as strong functional associations. No pathway–chemical relationship was directly verified at the biochemical reaction level. These findings support taxon-associated microbial functional assignments but do not establish enzymatic activity, metabolic flux, or direct microbial production of the measured metabolites or lipids.

**Supplementary Table S16. Targeted pathway–chemical association and functional-axis consolidation audit.**

Complete audit of targeted associations between prioritized microbial MetaCyc pathways and measured stool metabolites or lipids. The table includes longitudinal correlation measures, first-difference associations, and directional concordance across T4→T5, T5→T6, and T4→T6, together with chemical membership and functional-axis assignments. Positive and inverse relationships were considered according to biological expectations. Chemically indirect relationships were retained as functional associations or ecosystem-state markers rather than classified as directly verified biochemical mechanisms. The analysis was restricted to independently prioritized microbial pathways rather than indiscriminately testing all MetaCyc pathways against every measured chemical.

**Supplementary Table S17. Functional-axis associations with continuous longitudinal host immune features.**

Complete association audit between the five primary microbial–chemical functional axes and 65 continuous stool host-RNA immune features across T0–T9. The table retains longitudinal and first-difference correlations, transition-direction concordance, and associated evidence classifications. Relationships were evaluated without imposing a predetermined immune-response direction. The analysis supports the selected immune relationships shown in Figure 5 and the higher-resolution immune evidence summarized in Supplementary Data Figure S9. These associations are descriptive within a single individual and do not establish microbial causality or population-level statistical significance.

**Supplementary Table S18. Temporal-mechanistic consolidation of microbial functional, chemical, and immune-state transitions.**

Integrated longitudinal analysis comparing the T4→T5, T5→T6, and T4→T6 transitions across three biological stages: loss of the early microbial functional state, the *Bifidobacterium*-associated fermentation transition, and subsequent *Clostridium* lipid/CoA-associated activation. The workbook contains six worksheets: source and calculation documentation; stage-level transition summaries; complete T0–T9 stage trajectories; transition measurements for five primary functional axes; original axis trajectories; and transition measurements for all 65 continuous immune features. Standardized microbial functional, chemical-footprint, and selected immune-composite values are retained separately from calculated transition differences. The analysis supports the temporal succession model presented in Figure 5. Immune-composite values are preserved from the finalized source analysis; their precise feature membership and weighting should be documented in the associated computational provenance files.
